# Molecular epidemiology of HIV-1 in Oryol Oblast, Russia

**DOI:** 10.1101/2021.10.26.21265513

**Authors:** Ksenia R Safina, Yulia Sidorina, Natalya Efendieva, Elena Belonosova, Darya Saleeva, Alina Kirichenko, Dmitry Kireev, Vadim Pokrovsky, Georgii A. Bazykin

## Abstract

The HIV/AIDS epidemic in Russia is growing, with approximately 100,000 people infected annually. Molecular epidemiology can provide insight on the structure and dynamics of the epidemic. However, its applicability in Russia is limited by the weakness of genetic surveillance, as viral genetic data is only available for <1% of cases. Here, we provide a detailed description of the HIV-1 epidemic for one geographic region of Russia, Oryol Oblast, by collecting and sequencing viral samples from about a third of its HIV-positive population. We identify multiple introductions of HIV-1 into Oryol Oblast, resulting in 82 transmission lineages that together comprise 66% of the samples. Most introductions are of subtype A, the predominant HIV-1 subtype in Russia, followed by CRF63 and subtype B. Bayesian analysis estimates the effective reproduction number R_e_ for subtype A at 2.8 [1.7-4.4], in line with a growing epidemic. The frequency of CRF63 has been growing more rapidly, with the median R_e_ of 11.8 [4.6-28.7], in agreement with recent reports of this variant rising in frequency in some regions of Russia. In contrast to the patterns described previously in European and North American countries, we see no overrepresentation of males in transmission lineages; meanwhile, injecting drug users are overrepresented in transmission lineages. This likely reflects the structure of the HIV-1 epidemic in Russia dominated by heterosexual and, to smaller extent, IDU transmission. Samples attributed to MSM transmission are associated with subtype B and are less prevalent than expected from the male-to-female ratio for this subtype, suggesting underreporting of this transmission route. Together, our results provide a high-resolution description of the HIV-1 epidemic in Oryol Oblast, Russia, characterized by frequent interregional transmission, rapid growth of the epidemic and rapid displacement of subtype A with the recombinant CRF63 variant.

**Author Summary:** In 2017, Russia registered 138,843 new HIV-1 infections, which is nearly 5 times that of the second-highest European country. Meanwhile, genetic surveillance of HIV-1 in Russia is low, with only <1% of all HIV-positive samples sequenced. Here, we characterize the HIV-1 epidemic in Oryol Oblast, a region of Russia with an HIV-positive population of 2,157 as of 2019, by collecting viral genetic data covering a third of this population. We show that HIV-1 has been introduced into the region hundreds of times over the last 25 years, with many of the introductions resulting in sustained transmission within the region. By studying the branching patterns of the viral evolutionary trees, we reveal a persistently growing epidemic. A rapidly growing transmission cluster is associated with the CRF63 recombinant variant which has been spreading rapidly through the population of injecting drug users since its introduction around 2014. By analysing the subtype B which is characteristic of transmission between men who have sex with men (MSM), we show that the MSM transmission route is underreported. To our knowledge, this study is the most detailed description of the HIV-1 epidemic in a region of Russia.

## Introduction

HIV-1 poses a substantial threat to public health in Russia. Russia is characterized by one of the highest HIV-1 prevalence rates among European countries [1]. Since its initial introduction in the late 1980s, HIV-1 has been rapidly spreading across Russia due to wide-spread injecting drug usage and poor public awareness. In 2019, 97,176 new cases of HIV-1 were registered in Russia [2]. Its prevalence in the same year was estimated at 0,75% on the basis of registered cases [2]; the actual prevalence is likely higher. Currently, the epidemic predominantly develops through heterosexual transmission. In 2019, 63.9%, 33.0%, and 2.2% of all new cases with reported transmission route could be attributed to heterosexual, injecting drug-associated, and homosexual routes of transmission, respectively [2]. Low coverage of antiretroviral therapy (ARVT) contributes to poor epidemic control. In 2019, only 48.5% of the registered people living with HIV-1 in Russia were receiving therapy [3].

Analysis of molecular surveillance data can provide insights on pathways and rate of disease spread in an ongoing epidemic. However, the Russian diversity of HIV-1 remains poorly studied. A comprehensive description of the countrywide epidemic by methods of molecular epidemiology in Russia is hindered by the fact that genetic data on HIV-1 is available for less than 1% of the infected population. Coverage provided by molecular epidemiology studies of HIV-1 in specific regions of Russia is also low [4–8].

Here, we report a detailed molecular epidemiology analysis of the HIV-1 epidemic in a single region of Russia by covering a large fraction of its HIV-positive population. We focused on Oryol Oblast [9], a region with a population of 736,483 in the southwestern part of the Central Federal District of Russia [10]. As of 2019, there were 2,157 registered HIV-positive people in Oryol Oblast (Supplementary Fig. 1); we collect and analyse HIV-1 genetic data from 768 patients, thus covering over a third of the registered epidemic. Using a phylogenetic analysis, we infer 82 imports of HIV-1 into Oryol Oblast that were further transmitted within this region forming transmission lineages, as well as 250 imports that did not result in observed onward transmission, indicating unhindered spread between regions of Russia. Transmission lineages were enriched in injecting drug users but not in males, reflecting demographic properties of the epidemic. The epidemic is predominated by subtype A (87% of our dataset) followed by the recombinant CRF63 variant (7%). Using phylodynamic analysis, we show that subtype A is responsible for the moderate growth of the epidemic with R_e_ of 2.8 [1.7-4.4], while CRF63 demonstrates a much higher growth rate and should be closely monitored.

## Results

### The Oryol epidemic is largely constituted by the A subtype of HIV-1

We sequenced the *pol* region fragment of 858 HIV-1 samples obtained from 768 unique patients in Oryol Oblast (“Oryol dataset”). This dataset covers more than a third of the HIV-positive population in Oryol Oblast and represents an unbiased and well-annotated dataset (Fig. 1, Supplementary Fig. 1). Subtype composition in the Oryol dataset was more homogeneous compared to the non-Oryol Russian dataset represented by Genbank samples (“non-Oryol dataset”) (Fig. 1). Subtype A was the dominant subtype in Oryol Oblast (87.2%) recapitulating the historical Russian trend [11], followed by CRF63 (7.20%) and subtype B (2.49%). The transmission route was reported for 96% of the samples in the Oryol dataset, compared to less than 50% in the non-Oryol dataset; in both datasets, heterosexual transmission (HET) was the most prevalent, followed by transmission associated with injective drug users (IDU) and men who have sex with men (MSM), although the fraction of IDU and MSM samples was much higher in the non-Oryol samples. The male-to-female ratio was the same in both datasets (1.30), although sex has been reported for only 62% of non-Oryol samples. The fractions of sexes and reported transmission routes in the Oryol dataset were representative of those in the Oryol Oblast as a whole as reported by the Oryol AIDS center (Supplementary Fig. 1).

**Figure 1.**
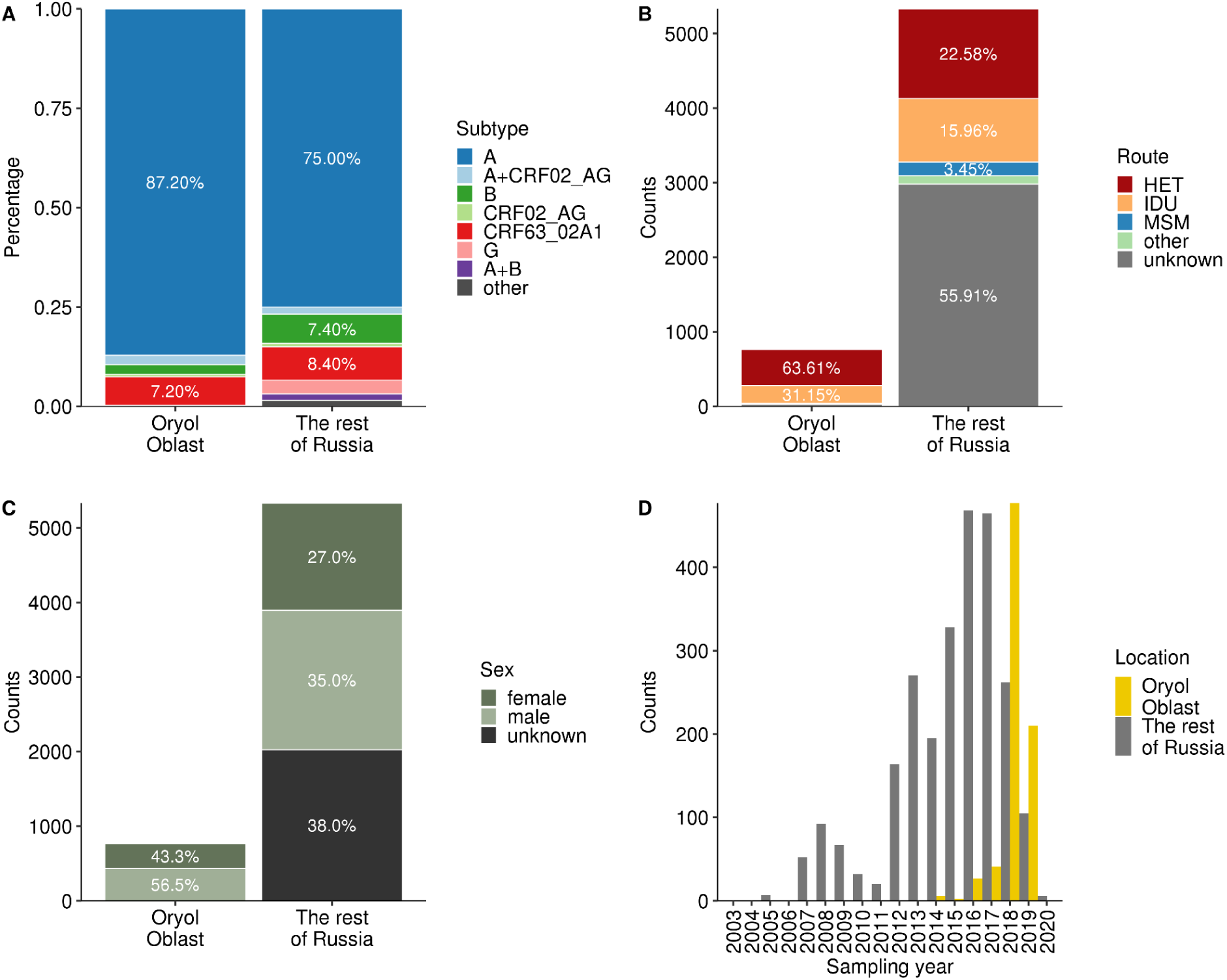
Statistics on the Oryol and non-Oryol datasets. The plots show the distribution of samples across subtypes (A), transmission routes (B), sexes (C), and sampling years (D). The seven subtypes most frequent in Russia are shown in A. Only sequences with complete sampling date are shown in D.

The phylogenetic tree reconstructed for the combined Oryol and non-Oryol dataset had separate clades corresponding to the three most abundant variants - subtypes A and B and variant CRF63, indicating that subtyping was mostly unambiguous (Fig. 2). In subsequent analysis, we focused on these three variants, covering a total of 739 Oryol Oblast samples from distinct patients.

**Figure 2.**
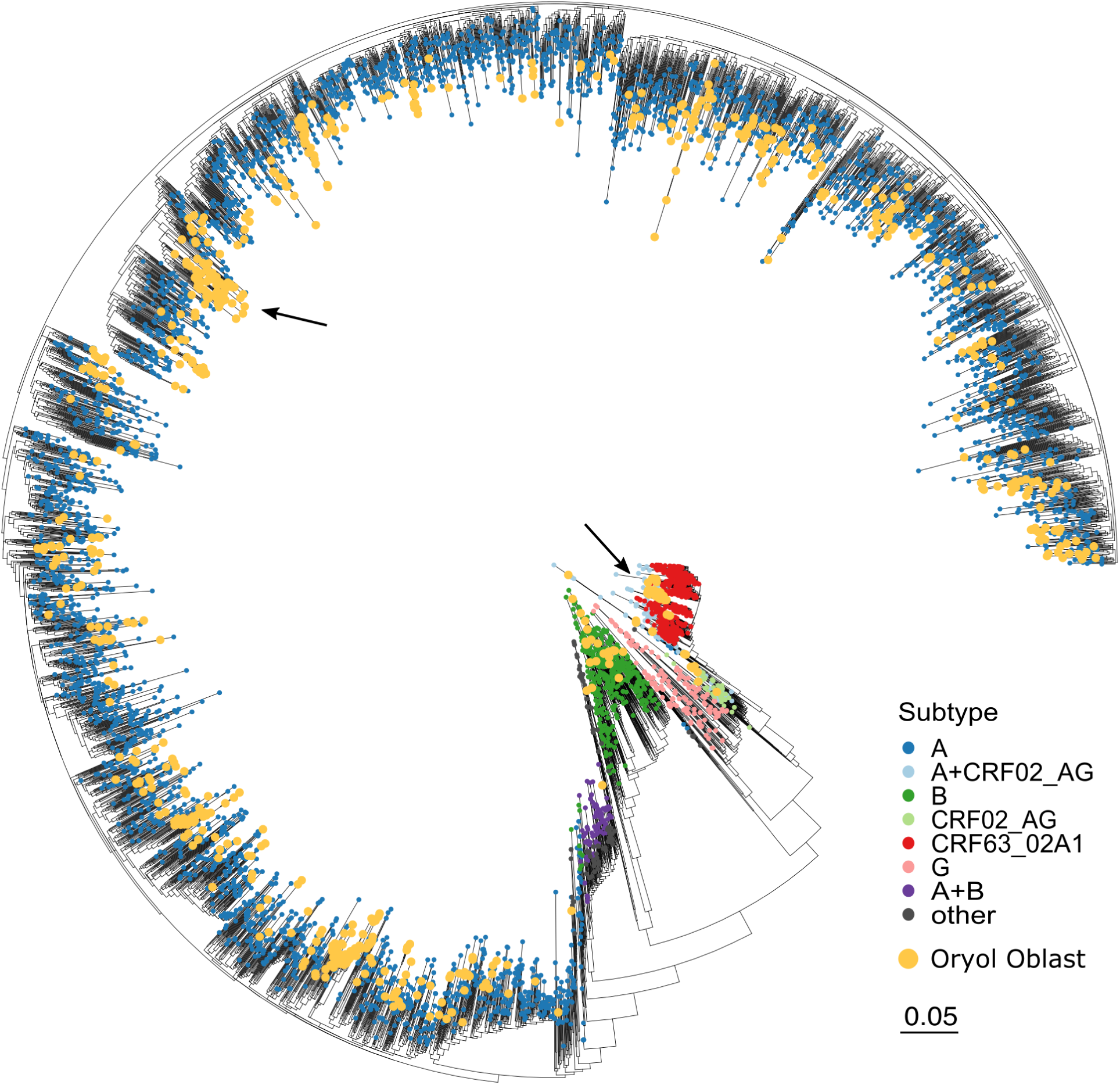
Phylogeny of the combined Russian HIV-1 dataset. When more than one sample per patient was available, only the earliest sample was used. Yellow dots mark Oryol Oblast samples. The two largest clades analysed separately, A and CRF63, are indicated with arrows. See Supplementary Fig. 2 for the phylogeny including all samples from repeatedly sequenced patients.

### HIV-1 has been imported into Oryol Oblast hundreds of times

To understand the interregional transmission routes of HIV-1, we first reconstructed separate phylogenetic trees for subtypes A and B and CRF63 (Supplementary Fig. 3). Overall, the Oryol dataset samples of each of the three variants were rather scattered across the Russian phylogeny (F_st_ = 0.01, 0.03, and 0.40 for variants A, B, and CRF63, respectively), supporting intensive transmission between regions. Still, many Oryol samples clustered on the phylogeny with other Oryol samples, indicating that many of the infections occurred within the region (Fig. 2, Supplementary Fig. 2,3).

We identified introductions of HIV-1 into the region as described in the Methods. We attributed 489 of the 739 Oryol samples to a total of 82 imports each resulting in one or more inferred transmissions within Oryol Oblast (“Oryol transmission lineages”). The remaining 250 sequences each resulted from its own import (“singletons”), for a total of 332 imports (Fig. 3). СRF63 was the most homogeneous variant with 94.5% (52/55) of the samples resulting from a single import, in agreement with its substantially higher F_st_ compared to subtypes A and B. We found seven non-Oryol samples descended from Oryol transmission lineages, indicating exports. Notably, the inferred numbers of imports and exports are not comparable: in particular, we likely undercount exports as the sampled diversity outside the region is low.

**Figure 3.**
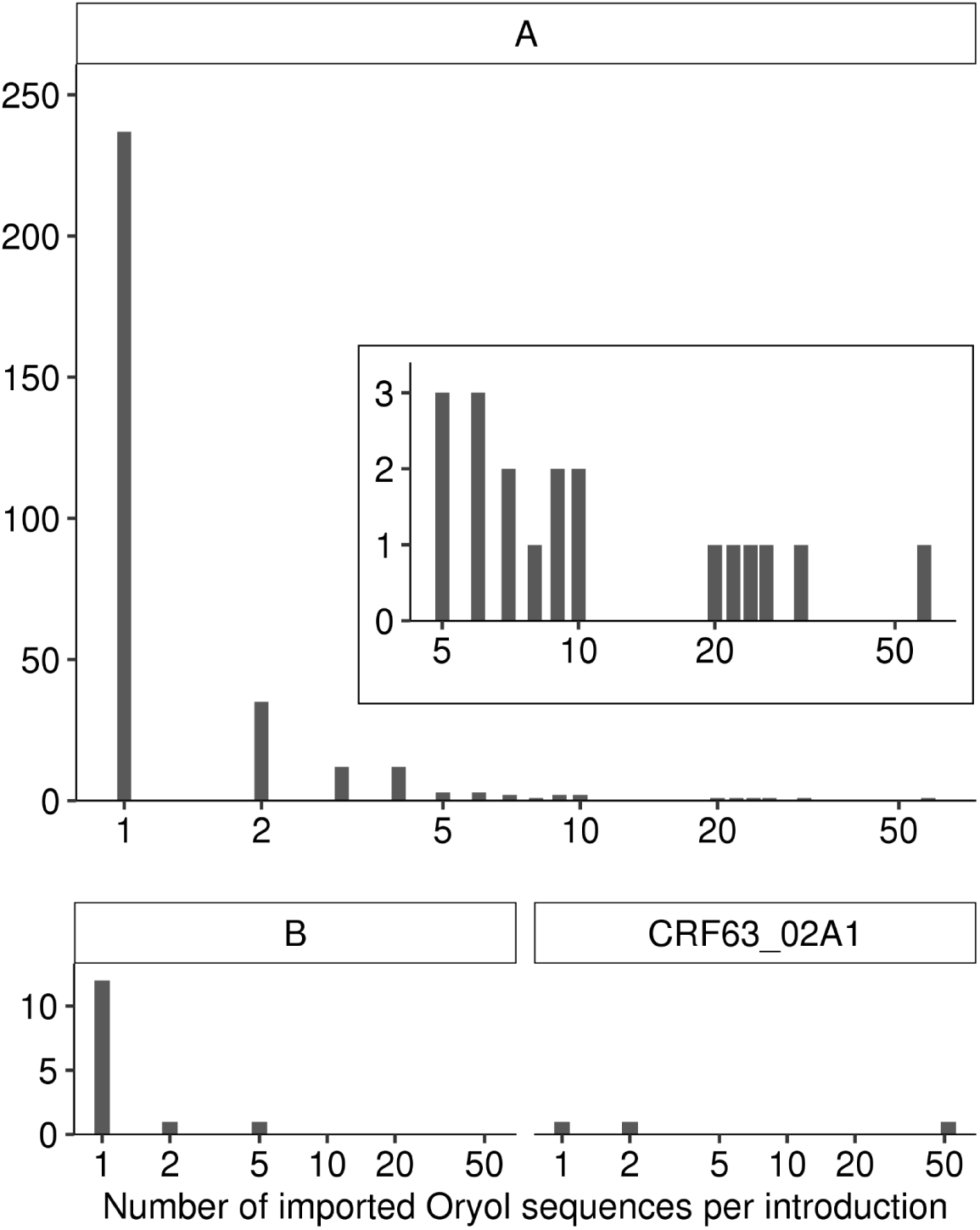
The number of Oryol sequences per import. The inset plot in A magnifies imports of subtype A of size 5 and more.

The number of imports is robust to the number of non-Oryol sequences available, already reaching a plateau when a comparable number of Oryol and non-Oryol sequences is used (Fig. 4A). This means that we estimate the minimal number of imports resulting in the sampled Oryol diversity robustly (at ∼332). Conversely, the number of imports depends strongly on the number of Oryol samples available (Fig. 4B).

**Figure 4.**
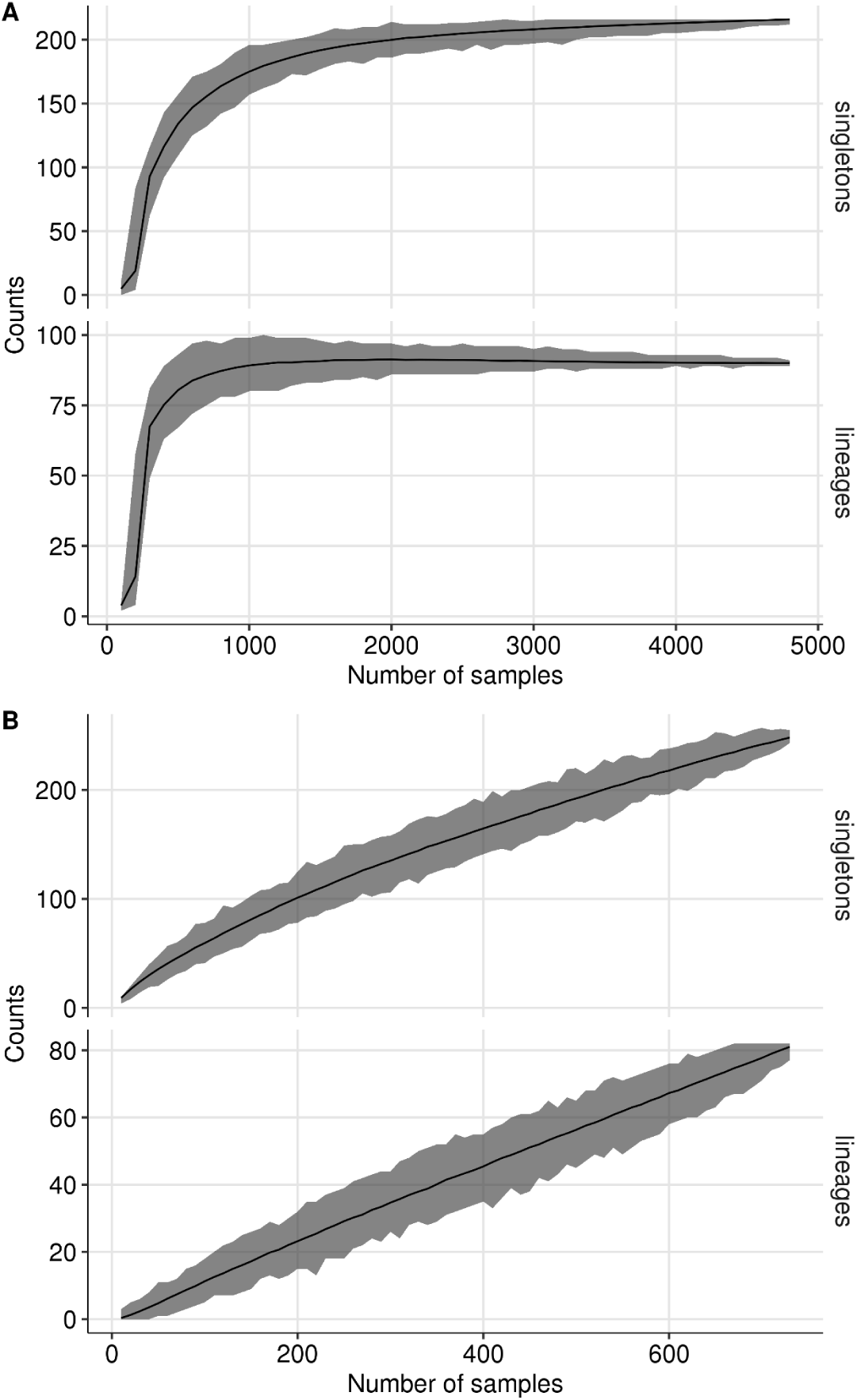
The dependence of the inferred number of singletons and transmission lineages on the number of non-Oryol (A) and Oryol (B) sequences used.

### Early imports disproportionally contributed to the epidemic in the region

To better understand the dynamics of transmission lineages, we dated the last common ancestor (LCA) of each lineage in subtype A and CRF63 as described in the Methods (Fig. 5). The reconstructed LCAs for individual lineages dated between 1996 and 2018, indicating that the genetic diversity within the currently sampled transmission lineages has accumulated over decades. On average, the first positive immunoblot for a lineage was obtained 0.91 years after this lineage was established based on the LCA date estimate, although the variance of this value was very high (Supplementary Fig. 6), in part due to lineages established after the first HIV-1 diagnosis in the lineage suggesting transmission of a non-basal variant. Lineages with earlier LCAs tended to have earlier first IB (Supplementary Fig. 7), validating this approach. Such lineages were also larger (linear regression p-value for the LCA date is 10^-4^; Fig. 7A), resulting in a disproportionate number of infections. Indeed, 50% of the lineages were established before May, 2010, but they were responsible for 70% of the observed cases (Supplementary Fig. 8).

**Figure 5.**
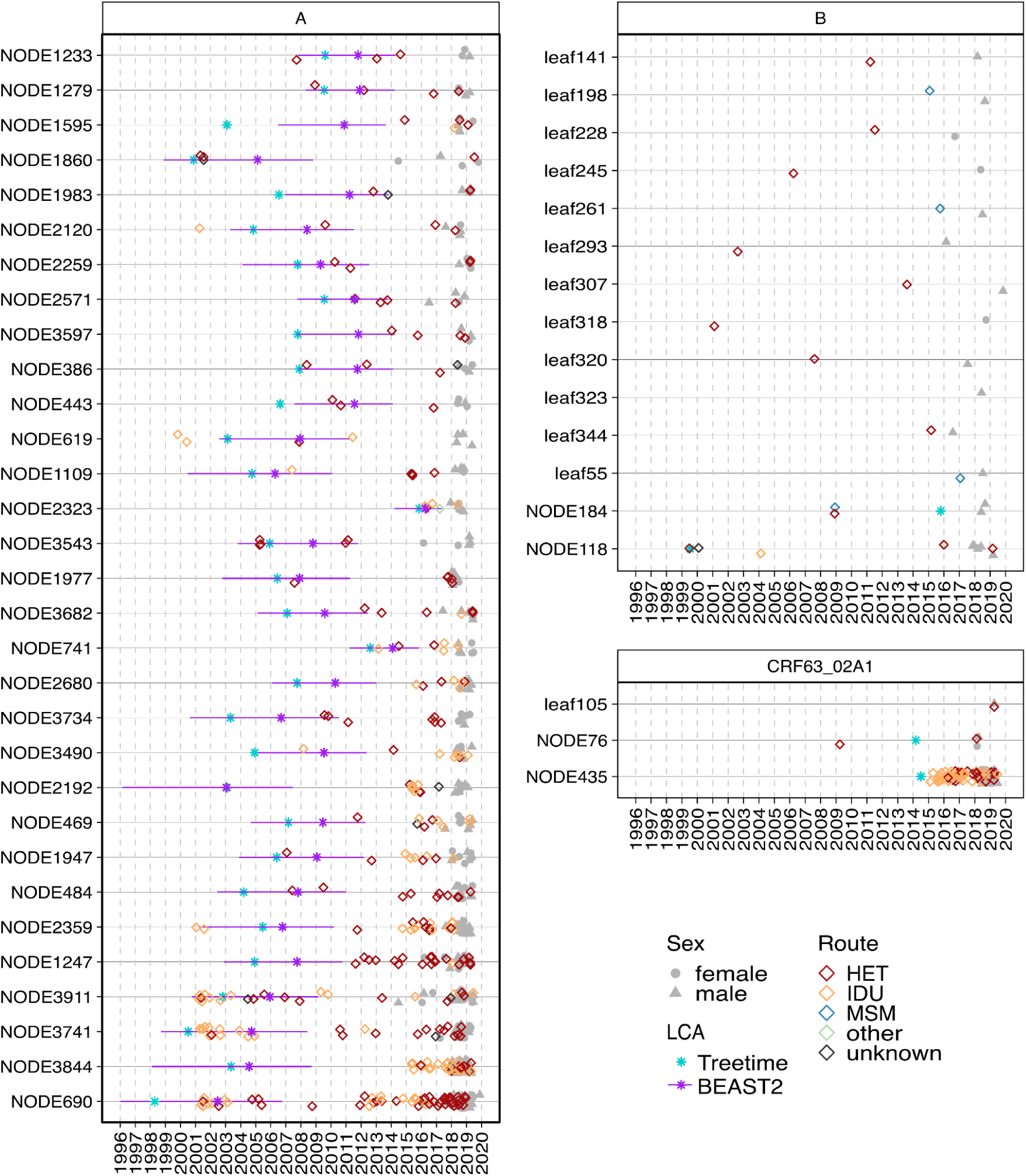
Temporal dynamics of Oryol HIV-1 lineages. Each horizontal line corresponds to a transmission lineage (with “NODE” prefix) or a singleton (with “leaf” prefix). Individual patients are shown twice: as an empty diamond at the date of diagnosis, with the color indicating the reported transmission route; and as a grey circle (for females) or triangle (for males) at the date of sampling. Only samples with complete sampling dates are shown. Cyan asterisks show the lineage LCA dates estimated by Treetime. For subtype A, purple asterisks and lines show the lineage LCA dates and 95% HPDs estimated by the multi-tree birth-death analysis. For subtype A, transmission lineages with at least four Oryol samples are shown. See Supplementary Fig. 4 for all subtype A lineages and singletons, and Supplementary Fig. 5 for all lineages and singletons sorted by IB date.

**Figure 6.**
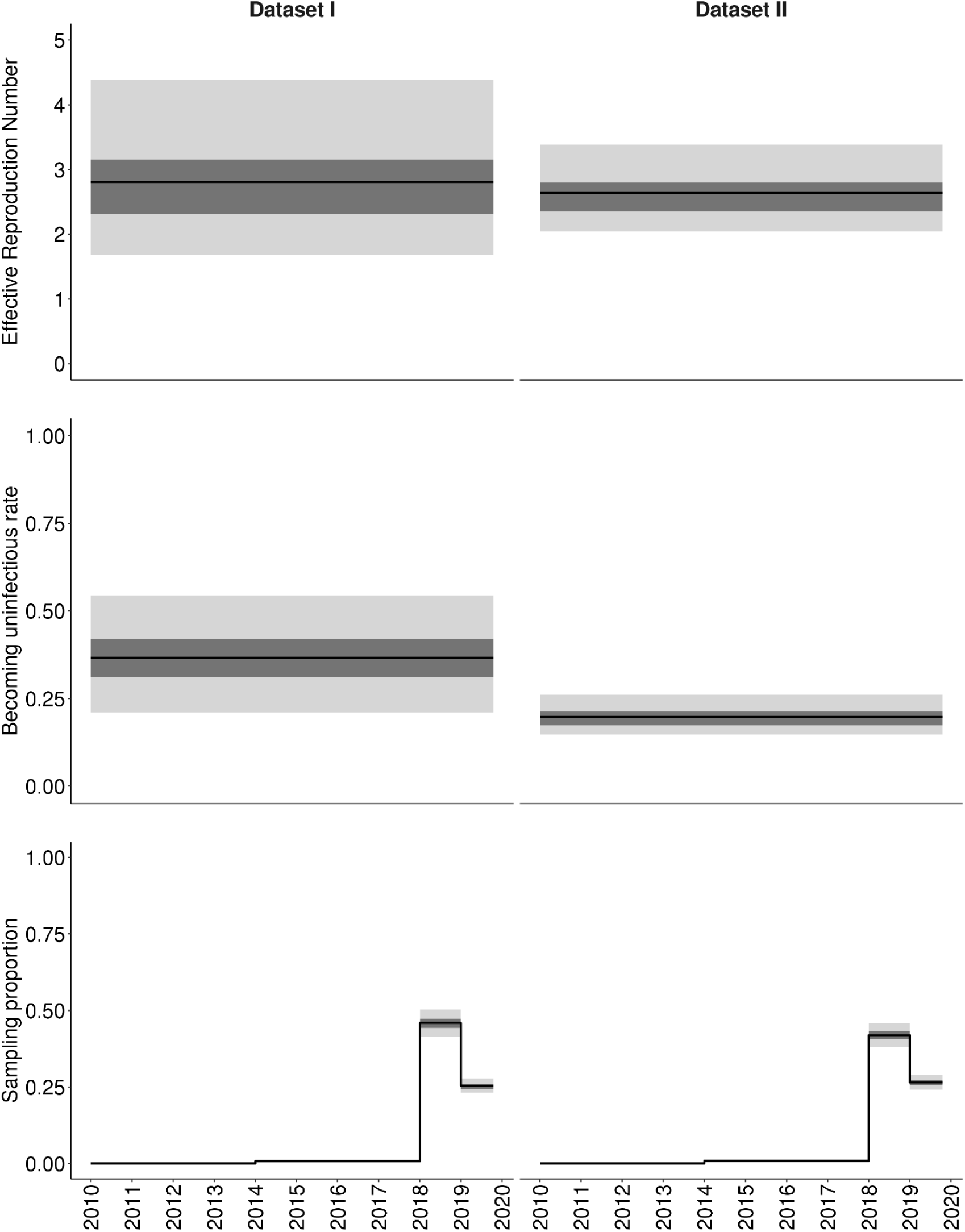
Epidemiological parameters inferred for the subtype A sub-epidemic. Dataset I, samples collected within a year of HIV-1 diagnosis; Dataset II, all samples. Solid line, median value; dark grey and light grey, 50% and 95% HPDs, respectively.

**Figure 7.**
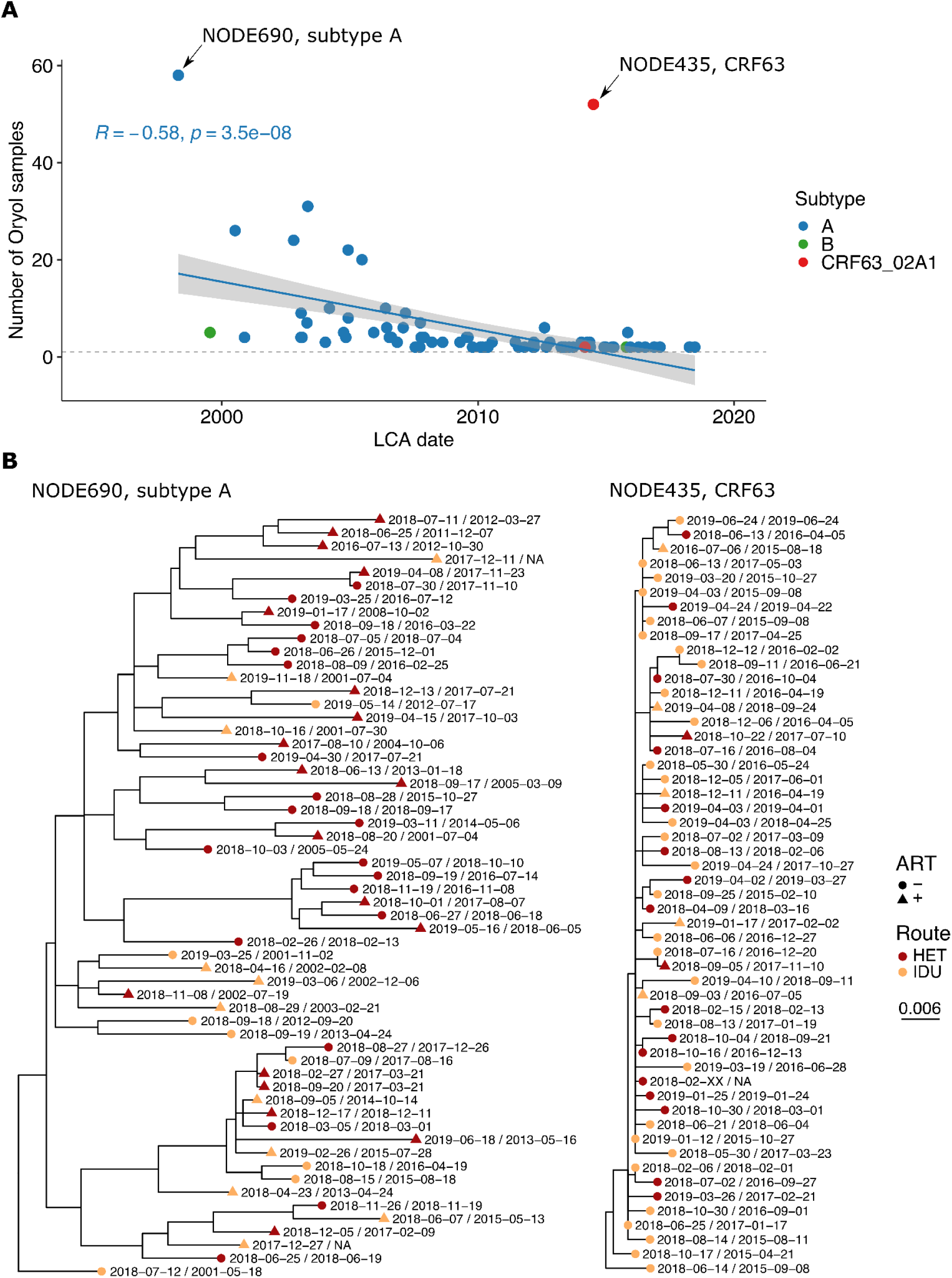
The comparison of the largest lineages of subtype A and CRF63. A. Lineages with earlier LCAs tend to carry more Oryol samples. The regression line is shown for subtype A; the regression analysis for all samples also shows a negative dependency (the slope p-value is 1e-04 for all lineages and 3.5e-08 for subtype A lineages). B. Maximum-likelihood phylogenies of the largest lineages of subtypes A and CRF63. Color indicates the reported transmission route, shape reflects the presence or absence of antiretroviral therapy at some point during the infection. The first and the second dates for each sample correspond to the date of sampling and the date of diagnosis, respectively.

### Epidemiological parameters of the subtype A sub-epidemic

Since subtype A represents 87% of all HIV-1 cases in Oryol Oblast in our dataset, we focused on this subtype to estimate the dynamics of the HIV-1 epidemic in this region. We used BEAST2 to infer epidemiological parameters of the subtype A sub-epidemic by simultaneously analysing all transmission lineages and singletons of subtype A. The recently developed multi-tree implementation of BEAST2 [12] allows treating separate lineages as realizations of the same epidemiological process whose parameters can be jointly inferred. We modeled the subtype A sub-epidemic as a birth-death process with a constant time-independent reproductive number, constant rate of becoming noninfectious, and time-dependent multidimensional sampling proportion upon which we put a strong prior (see Methods). The prior on the sampling proportion was estimated based on a set of samples that were sequenced less than a year after the initial diagnosis (denoted as ‘Dataset I’ in Fig. 6). We used this prior both for the dataset comprising only early infections (Dataset I) and for the full set of samples (Dataset II); the results did not differ drastically (Fig. 6, left vs. right column). The median effective reproductive number (R_e_) was 2.8 and 2.6 for datasets I and II, respectively; the corresponding rates of becoming uninfectious were 0.37 and 0.20. The 95% HPD for R_e_ inferred from the full Dataset II is strictly higher than 2, implying a growing epidemic. R_e_ estimated from the reported yearly incidence by EpiEstim is also above 1, although it is lower than that obtained using the birth-death model (Supplementary Fig. 14).

### Bayesian phylogenetic analysis indicates rapid growth of the CRF63 lineage in Oryol Oblast

The major CRF63 lineage (NODE435 in Fig. 5) is largely transmitted through IDU (IDU is the reported transmission route for 66% of the cases, compared to 31% of the epidemic as a whole; Fig. 5). This lineage is unexpectedly large for its age (Fig. 7A), indicating rapid spread through the IDU population. Indeed, in less than ten years, it has reached the same size as the largest lineage of subtype A (NODE690 in Fig. 5) which has circulated for at least two decades. Consistently, the phylogeny of the CRF63 samples is characterized by a more recent LCA and a higher fraction of multiple merger events compared to the phylogeny of the A samples obtained at similar times (Fig. 7B), suggesting a more rapid spread of CRF63. To test whether the two lineages indeed differ in their dynamics, we used two approaches. First, we utilized the coalescent approach to infer the growth rate of both lineages assuming logistic growth (Fig. 8A, Supplementary Fig. 9). Second, we used the BDSKY model to directly infer epidemiological parameters, i.e. the reproduction number and the rate of becoming uninfectious (Fig. 8B). Importantly, for the largest clade of subtype A, BDSKY produced R_e_ estimates similar to those of a multi-tree BEAST2 implementation for multiple introductions of A (Fig. 6), indicating the robustness of R_e_ estimates. Both the coalescent and the BDSKY models suggest a higher growth rate of the CRF63 lineage compared to the largest lineage of subtype A (Fig. 8).

**Figure 8.**
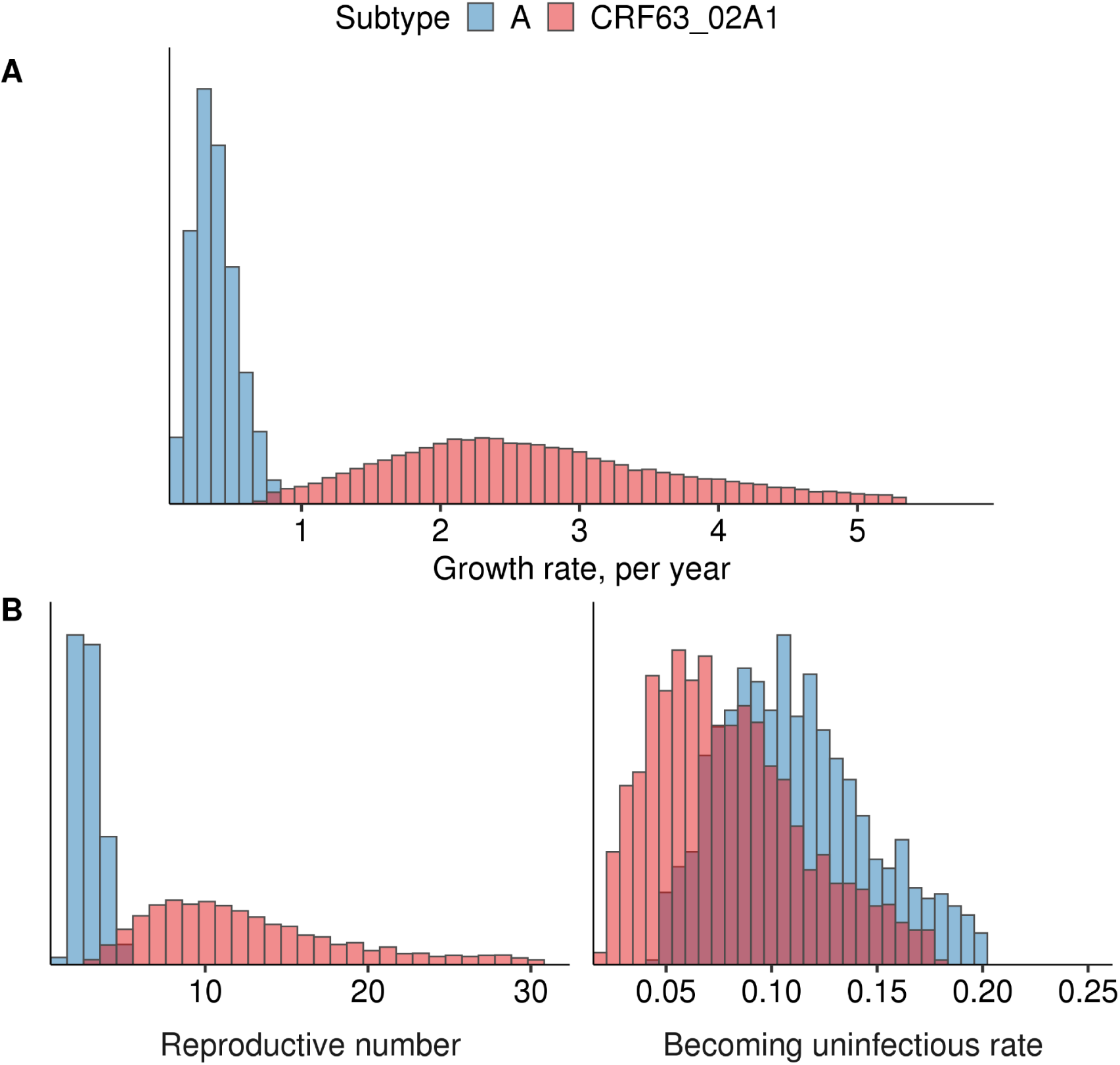
Phylodynamic inferences for the two transmission lineages shown on Fig. 7. A. 95% HPD of the growth rate parameter in the coalescent logistic growth model. B. 95% HPD of the reproductive number and the rate of becoming uninfectious inferred by the BDSKY model.

### No evidence for the preferred mechanism of transmission at the origin of lineages

In the late 1990s to early 2000s, most newly reported HIV-1 infections were among IDUs (Supplementary Fig. 10). Consistently, we find that most of the early lineages were first sampled in IDUs (Supplementary Fig. 11), suggesting that they were founded by them. However, we see no evidence for IDUs being more likely to be the originators of lineages when we control for the dates of lineage origins. Indeed, in the controlled matched-pair datasets, there is no difference in time from LCA to the earliest diagnosis between IDU- and HET- founded clusters, nor is there a preference in transmission route of the earliest sample in the lineage (Supplementary Fig. 12). This means that the high prevalence of IDUs in lineage origin reflects the temporal shift in the outbreak composition in our data rather than higher inherent spreading by IDUs.

### Distribution of gender and transmission route categories across import lineages

We next studied whether transmission lineages in subtype A are associated with transmission route and/or sex categories by comparing lineages and singletons (see Methods). We did not observe any overrepresentation of males within lineages (Fig. 9A) as described previously in other countries [13–16]. However, IDUs were overrepresented within lineages, with more samples belonging to lineages than expected by chance (Fig. 9B). Furthermore, there were fewer lineages carrying IDUs than expected randomly (Fig. 9D); in other words, an IDU was more likely to fall into a lineage with another IDU (p = 0.0007). No such difference was observed for males (Fig. 9C, p = 0.9923).

**Figure 9.**
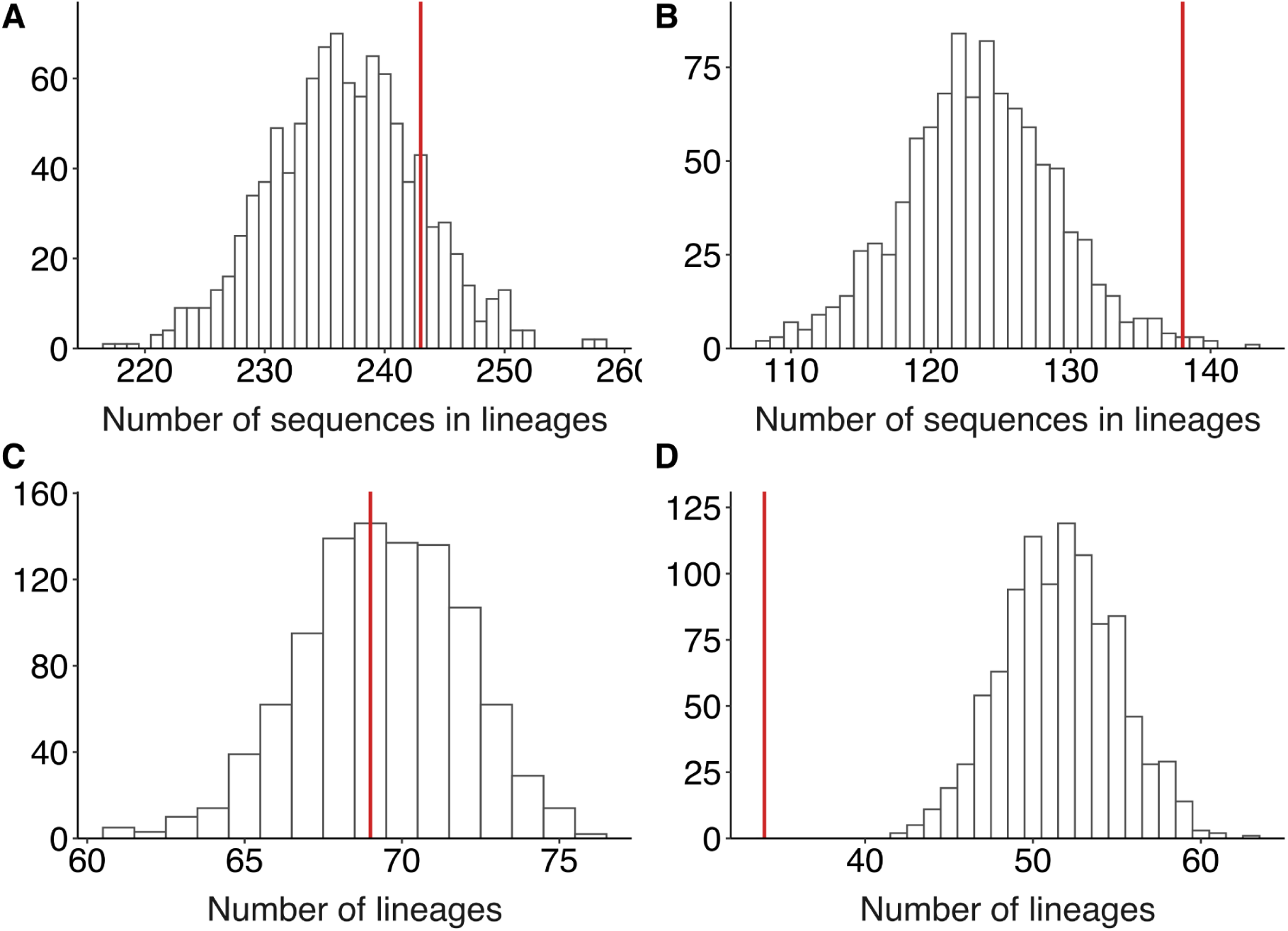
IDUs, but not males, are overrepresented within lineages. The expected distribution of the number of clustered sequences and the number of clusters carrying (A, C) males or (B, D) IDUs within subtype A. For variants B and CRF63, see Supplementary Fig. 13.

### MSM transmission route appears to be underreported in Oryol Oblast

The lack of overrepresentation of males in transmission lineages is perhaps unsurprising given the heterosexual nature of the Russian HIV-1 epidemic. Still, we might expect an excess of MSM reported transmission in subtype B which is mainly associated with the MSM transmission route worldwide [17] and in Russia [18, 19]. Indeed, the male-to-female ratio in subtype B is much higher than that in subtype A where we don’t expect an MSM-associated bias (16/3 vs. 369/296, Fisher’s exact test p=0.0169), and subtype B carries 5 out of 6 MSM-associated samples in our dataset (Fig. 10).

**Figure 10.**
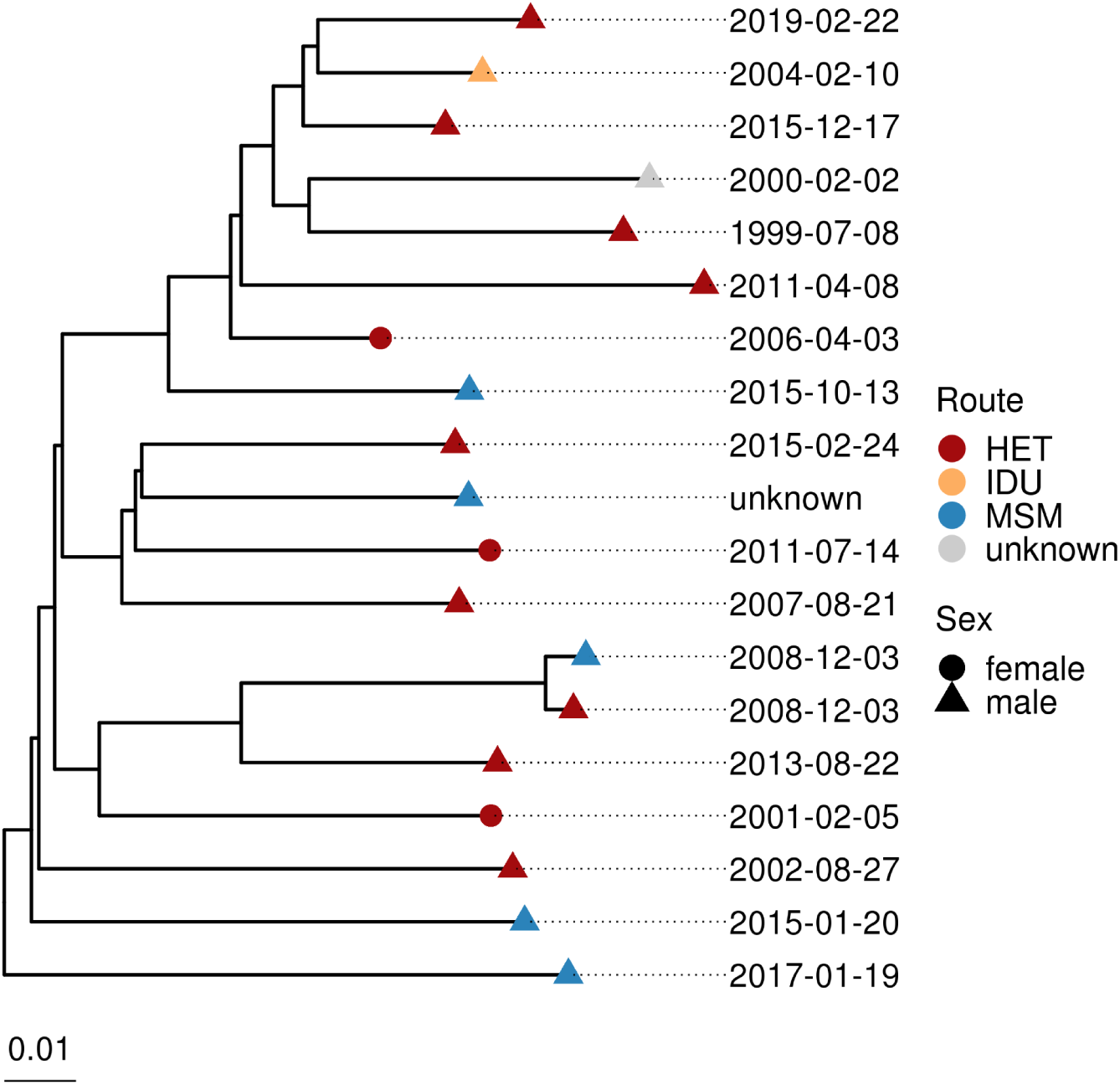
Phylogenetic tree of subtype B in Oryol Oblast. HIV-1 diagnosis dates are shown at tips.

Is MSM transmission adequately reported in our data? Underreporting of this transmission route can be estimated from the sex ratio among those samples for which other transmission routes are reported. Among the 14 subtype B samples with the non-MSM reported transmission route, only 3 came from females. Based on the sex ratio in subtype A, the number of males expected from this number of females is ∼4. In fact, however, 11 males are observed. While the difference from the expected sex ratio is not statistically significant (Fisher’s exact test, p=0.1054), if confirmed, such an excess of males would correspond to a ∼2.4-fold underreporting of the MSM transmission route.

## Discussion

Since its beginning in the 1980s, the HIV-1 epidemic in Russia has been growing due to poor public awareness, stigmatization of the key risk groups, and insufficient funding [20–23]. Molecular epidemiology is informative of the characteristics of the epidemic, such as its genetic composition and reproductive number; it can also shed light on details of individual outbreaks, e.g. by identifying an increased transmission risk among a certain group or revealing a rapidly growing transmission cluster. However, molecular epidemiology methods work poorly when sampling is low, which is the case for the Russian epidemic where less than 1% of reported cases is accompanied by genetic data. Moreover, HIV-1 genetic data available in Genbank is partially obtained through target studies focusing on specific transmission routes [7,18,24,25] or outbreaks [26], making it unrepresentative of the epidemic as a whole.

Unfortunately, given the magnitude of the HIV-1 epidemic in Russia, it is infeasible to rapidly achieve sufficient genetic coverage for the entire country. Here, we instead considered a single geographic region of Russia. We focused on Oryol Oblast, a relatively small region of Russia with a total population of 736,483 [27] and a registered HIV-positive population of 2,157 (Supplementary Fig. 1) as of 2019. The registered HIV-1 incidence is lower than that in Russia as a whole (0.29% vs. 0.73%). The relatively small size of the HIV-positive population allowed us to attain representative coverage of the local epidemic.

The dataset analysed in this work comprises sequences collected from 768 patients, or 36% of the registered HIV-positive population. Compared to non-Oryol Russian sequences available from Genbank, the Oryol dataset is unbiased, better annotated, and more up-to-date (Fig. 1). It also agrees well with the official sequence-independent statistics reported by the Oryol Regional Center for AIDS (Supplementary Fig. 1). The fraction of IDUs in our dataset is slightly below that in the official statistics (31.2% vs. 37.0%) which might be explained by lower adherence to AIDS center visits and/or lower survival among the members of this risk group [28]. In line with the official statistics, our dataset also captures the change in the predominant transmission route from IDU to heterosexual transmission in the 2000s, followed by an abrupt increase in the fraction of IDUs since 2014 when a novel designer drug became widespread [29] (Supplementary Fig. 1,10). The male-to-female ratio is also similar to that in the official statistics (1.7 vs. 1.5) and is close to the country-wide ratio of 1.6 (as of 31.12.2019; [2]).

The subtype composition in Oryol Oblast is more homogeneous compared to non-Oryol Genbank (Fig. 1A), due to interregional differences and/or targeting of specific subtypes in previous studies. Still, both in Oryol Oblast and in Russia in general, the most abundant subtype is A which has historically dominated the HIV-1 epidemic in Russia [11]. Subtype A is genetically diverse and divided into several clades, or sub-subtypes. Initially, Russian sequences belonging to subtype A were annotated as A1, the most widespread subtype A clade. Recently, however, this ‘Russian A1’ was demonstrated to be genetically different from the African A1 [30] and received its own identifier A6. We kept the broader ‘A’ identifier in the text for consistency with the results produced by the SierraPy subtyping tool that we used; another widely used subtyping tool that we checked, REGA [31], utilizes an old set of reference sequences and still annotates the Russian subtype A as A1.

For the three most abundant variants (A, B, and CRF63), we used the reconstructed maximum likelihood phylogenetic trees to infer imports into Oryol Oblast. Our approach is based on the assumption that all imports are of Russian origin. While some imports could also directly come from other countries, such cases are probably rare as transborder travel is expected to be much less intense than within-country travel.

The inferred number of transmission lineages was robust to the number of non-Oryol sequences used, meaning that we have successfully resolved the sequenced genetic diversity of HIV-1 in Oryol Oblast (Fig. 4A). However, this number was strongly dependent on the amount of Oryol samples available: it did not saturate as we increased the number of Oryol samples in the analysis (Fig. 4B). This implies that the diversity of HIV-1 in Oryol oblast is higher than can be captured by sequencing of a third of the population, and/or that there is a constant import of novel HIV-1 variants into the region.

Lineages that were established early contained more samples, implying that early imports into the region significantly contributed to the current epidemic. Early imports were mostly associated with IDUs; however, we did not find evidence for preferred seeding of lineages by IDUs (Supplementary Fig. 12), suggesting that the prevalence of different transmission routes was shaped by social factors rather than the biology of transmission.

Nearly two-thirds of all samples were attributed to transmission lineages. The remaining “singleton” sequences did not result in observable transmission within Oryol. A fraction of singletons could actually correspond to non-Oryol residents. Indeed, in Russia, the HIV data collected by an AIDS center is household based, meaning that patients are assigned to centers on the basis of their household registration. The attribution of sequences comprising Oryol transmission lineages as actually coming from Oryol Oblast residents is probably more reliable.

Using phylodynamic approaches, we characterized the epidemiological parameters of HIV-1 spread. These analyses have several limitations. First, we assumed that the evolutionary rate is independent of the age of the infection and the presence of therapy. Differences in infection duration and therapy between patients could have affected the estimated LCA dates. For example, the rate of evolution can be reduced by antiretroviral therapy [32], pushing the LCA estimates to the present. Such biases could lead to some unexpected LCA datings that we observe. For example, in several instances (e.g. NODE619 and NODE2359 in subtype A or NODE184 in subtype B), two or more of the earliest diagnoses in a transmission lineage had earlier dates than the reconstructed LCA of this lineage, which is impossible as LCA by definition must correspond to the earliest transmission between samples of the lineage. We speculate that this discrepancy could have been caused by differences in the antiretroviral therapy status between sampled individuals; indeed, while ∼40% of our dataset was on therapy, one of the two earliest samples in both NODE619 and NODE2359 lineages (of subtype A) and both patients in the NODE184 lineage (of subtype B) receive(d) therapy (Fig. 5). In theory, it may be possible to account for differences between patients in viral evolution rates, e.g. by inferring two distinct evolutionary rates, but the amount of variation in duration and adherence to therapy would be hard to account for. Overall, our results seem qualitatively robust to the choice of evolutionary rate.

Second, birth-death models are unidentifiable unless at least one of the parameters is fixed or strongly constrained. We put a strict prior on the sampling proportion, defined as the fraction of all infections being sampled. Sampling density was strongly non-uniform across years, with the vast majority of samples collected over just two years (2018-2019); however, many of these patients became infected years ago. We thus constructed a four-dimensional prior on sampling proportion based on the subset of “rapidly sequenced” samples (Dataset I, see Methods), and used it for datasets that also included old infections and for which we could not make an informative assumption about sampling. While the parameters estimated from different datasets were similar (Fig. 6), those estimated on the basis of Dataset I should probably be considered more reliable.

Third, birth-death models assume that sampling of an infection results in its “death” due to host recovery or change of behaviour. Unfortunately, this assumption usually does not hold for long-term infectious diseases such as HIV-1, especially in countries like Russia where treatment is available to less than 50% of HIV-positive people [3]. This may make our inferences about the reproduction number and the rate of becoming uninfectious under- and overestimated, respectively.

Fourth, the obtained phylodynamic estimates are relevant for the identified part of the Oryol Oblast epidemic. The unidentified part of the epidemic, i.e., associated with non-registered cases (which may comprise up to 20% in Russia [33]), can grow more rapidly due to lack of awareness of the HIV-1 diagnosis [34] and a definite absence of therapy among these people.

Fifth, our analyses in the main text use all sites, including those of drug resistance mutations (DRMs). Changes at such sites are frequently recurrent between patients, and therefore may obscure phylogenetic analyses. Although drug-resistance mutations were reported not to bias the composition of transmission lineages [35], they were shown to affect some analyses, e.g. the exact reconstructed history of transmissions [36]. To address this, we repeated the maximum-likelihood tree reconstruction, definition of transmission lineages (Supplementary Fig. 15), the birth-death analysis on subtype A (Supplementary Table 5), and the comparison of the two largest clades in variants A and CRF63 analyses (Supplementary Fig. 16) on DRM-masked alignments. We found that inclusion of DRM sites did not affect our conclusions.

With these limitations in mind, we assessed the characteristics of the epidemic. As subtype A currently prevails in Oryol Oblast, epidemiological parameters of its sub-epidemic should tone the dynamics of HIV-1 in the region. We jointly inferred R_e_ and the rate of becoming uninfectious for all imports of subtype A. The estimates produced from both Dataset I and Dataset II covering subtype A sub-epidemic consistently inferred R_e_ above 1 (median values are 2.8 and 2.6, Fig. 6), implying a growing epidemic. For the above-mentioned reasons, these values should be considered as lower boundaries; the actual growth rate is probably higher. The rate of becoming uninfectious is estimated as 0.37 and 0.20 for datasets I and II, which is equivalent to the total duration of infection of 2.7 and 5 years, respectively.

Independently, we inferred the R_e_ dynamics from case count data using EpiEstim (Supplementary Fig. 14). These estimates of R_e_ were lower, compared to the birth-death model, although still strictly higher than 1. There can be at least two reasons for this discrepancy. First, reliable incidence data is only available starting from 2000, and the sliding window approach implemented in EpiEstim does not allow us to obtain R_e_ estimates for years before 2005. If R_e_ in 1990s and early 2000s was in fact higher than that later in the epidemic, the EpiEstim estimate would not be reflective of the epidemic as a whole, and would be an underestimate. Second, R_e_ can be biased by heterogeneity in sampling procedure; for instance, if the epidemic shifts to the heterosexual population but infected heterosexuals are diagnosed more slowly, the count-based R_e_ estimate would be lowered.

Besides subtype A, we separately studied CRF63 which recently evolved in the Siberian part of Russia as a result of recombination between CRF02 and subtype A6 and has been spreading across the regions of Russia since then. We show that in Oryol Oblast, it is mostly represented by a single introduction event that resulted in 52 identified infections. This transmission lineage was unexpectedly young for its size, motivating us to compare epidemiological dynamics of subtype A and CRF63. We compared the two largest clades of these variants using two phylodynamic approaches. First, we inferred the growth rate of both clades assuming logistic growth; second, we inferred the reproduction number and the rate of becoming uninfectious using a birth-death model and a sampling prior used for multi-tree subtype A analysis. Both analyses indicate a higher growth rate of the CRF63 clade.

The rapid invasion of CRF63 in Oryol Oblast is consistent with the fact that this variant has been rapidly expanding in several Russian regions in recent years, and has become the dominant variant among new infections in some of them [37–39]. The differences in the rate of spread of A and CRF63 could result from biological and/or epidemiological differences between these two variants. CRF63 has originated from recombination between CRF02 and the major Russian strain A6 [40], and its success could be due to difference in properties between CRF02 ([41, 42], but see [43]) and A6. Indeed, while the biological properties of CRF63 have not yet been studied extensively, this strain has been reported to be antigenically distinct from A6 [44]. Further experimental studies are required to test the possible biological differences between CRF63 and A6.

Nevertheless, the rapid growth of CRF63 is probably more likely to result from its characteristic epidemiology. First and foremost, the CRF63 clade carries a much higher fraction of IDUs. Second, a smaller fraction of CRF63-infected patients were reported to receive therapy (16% vs. 40% in our dataset, Fig. 7). Both these factors facilitate transmission. Third, as CRF63 was introduced into Oryol Oblast much later than subtype A, infections associated with it are expected to be younger even when controlling for the year of diagnosis, and younger infections are associated with higher transmission [45]. Overall, the rapidly growing CRF63 transmission cluster in Oryol Oblast merits close monitoring.

Outside CRF63, the distribution of transmission routes is also informative about the structure of the epidemic. Within the predominant subtype A, we did not observe excessive clustering among males such as has been reported in many countries where the MSM-associated subtype B is prevalent [13–16]. This might be explained by a different structure of the HIV-1 epidemic in Russia. Compared to those areas where most new diagnoses come from MSM contacts resulting in a majority of the HIV-positive population being male, HIV-1 in Russia heavily affects the general population through HET contacts as well as the IDUs, making the male-to-female ratio less biased (Fig. 1, refs on official stats on Russia).

By contrast, we do observe a clustering of IDUs in transmission lineages of subtype A. Part of this effect could arise from a possible fraction of non-Oryol residents outside transmission lineages (see above), e.g. if non-Oryol residents are less likely to be IDUs. However, we also observe that IDUs are more likely to co-occur within the same transmission lineages, and this bias cannot have a purely geographic nature. Therefore, the observed IDU clustering probably results from increased transmission within enclosed communities of drug users, although this partially may also come from IDU being the major transmission route in early years of epidemic in Russia.

The MSM transmission route was rarely (0.8%) reported in our dataset. Still, five of six MSM cases belong to subtype B in agreement with the historical association of subtype B with this route of transmission. The prevalence of males with non-MSM reported transmission route in subtype B suggests underreporting of MSM in this subtype.

In our dataset, only 39% of patients were on therapy; this fraction is lower than that reported by the Oryol AIDS center (65%). This is because a significant fraction of our dataset corresponds to recently diagnosed patients who have not started receiving therapy yet. Prior to 2018, according to government regulations, therapy was only provided to patients with CD4 counts below 500 [46]. While post-2018 recommendations propose therapy for all patients [47], and the fraction of HIV-positive people on therapy was increasing both in Oryol Oblast and in Russia in general, the funding allocated in 2019 [48] and in 2020 [49] only covered therapy for 43% and 46% of HIV-positive patients. Our finding of large R_e_ is in line with insufficient effectiveness of the policies currently in place to curb the HIV-1 epidemic in Russia. 464.138 505.190

Taken together, the results presented in this work offer the most in-depth molecular-based characteristic of the HIV-1 sub-epidemic within Russia. Multiple lines of evidence indicate that the HIV-1 epidemic in Oryol Oblast is clearly growing, mainly due to subtype A. We provide evidence that the frequency of the recently introduced CRF63 grows faster compared to subtype A, and suggest that this variant may eventually start to contribute more significantly to the Oryol epidemic. CRF63 should be a high-priority target for molecular surveillance and experimental studies in Oryol Oblast.

## Methods

### Data collection and ethics

Patients were enrolled into the study between January 1, 2018 and June 30, 2019. Over this period, 681 blood samples were collected from HIV-infected people living in the Oryol city and the remainder of Oryol Oblast (Fig. 1) by the local AIDS center through a routine surveillance program and through regular check-ups of the registered HIV-infected people. Additionally, we included the 241 samples obtained between March 31, 2014 and November 2, 2019 in the course of a study on drug resistance, for a total of 922 samples from Oryol Oblast. HIV-1 RNA for sequencing was obtained from blood plasma left after the viral load analysis. Demographic, clinical, and epidemiological data for participants were obtained from their medical records. The assumed route of infection was recorded by interviewing the patients. Written informed consent was obtained from all subjects. The study was approved by the Local Ethics Committee of the Central Research Institute of Epidemiology (protocol 93).

### Sequencing

Sequencing was performed between June 29, 2019 and March 30, 2021. The sequence of the *pol* region covering the protease gene and part of the reverse transcriptase gene was obtained either by Sanger or by next-generation sequencing (NGS). In both cases, RNA was isolated from blood plasma using phenol chloroform extraction. For Sanger sequencing, the AmpliSens HIV-Resist-Seq (CRIE, Russia) kit for in vitro diagnostics was used according to the manufacturer’s instructions. For NGS, a two-step amplification procedure was used. The first step of amplification was combined with reverse transcription. Amplification was performed according to the following protocol: 45°C - 30 min; 95°C - 15 min; 30 cycles: 95°C - 30 s, 50°C - 30 s, 72°C - 1 min 30 s; 72°C - 5 min. During this stage, a DNA fragment of approximately 1.5 kb was amplified (positions 2074-3539 in the reference HIV-1 strain HXB2, GenBank K03455). The second step of amplification was performed in four independent tubes for each sample. Amplification produced four overlapping DNA amplicons that ranged in size from 427 to 586 bp. This approach made it possible to simplify library preparation and eliminated the need for DNA fragmentation. After purification with Sera-Mag Magnetic Speedbeads (GE Healthcare Biosciences) magnetic particles, the amplified fragments were mixed in equal proportions. After barcoding, next-generation sequencing was performed on Illumina Miseq machine (Illumina, USA) with the MiSeq Reagent Kit V3 (600 cycles). Totally, out of the 681 samples collected in this study, 562 samples were sequenced using NGS, and the remaining 119, using Sanger technology.

### Iterative consensus calling

The diversity of Russian HIV-1 differs significantly from the widely used HXB2 reference, complicating variant calling. Furthermore, amplification of the pol fragment via four slightly overlapping amplicons prevented the use of de novo assembly tools like IVA [50]. To address these issues, we developed the following custom consensus calling pipeline, and applied it to each of the 562 samples sequenced on Illumina platform:

1. Trim paired reads using Trimmomatic-0.33 [51] (with options ILLUMINACLIP:$adapters:2:30:10 LEADING:5 TRAILING:5 SLIDINGWINDOW:5:15 MINLEN:50);
2. Align the trimmed reads against a set of curated reference sequences from LANL HIV (198 seqs, [52]) using blastn v.2.2.31 [53] with default options; select the closest reference sequence REF_DRAFT;
3. Transfer the coordinates of the four pairs of primers from the HXB2 reference to the selected REF_DRAFT; Assign REF_INIT = REF_DRAFT;
4. Map the trimmed reads against REF_INIT using smalt v.0.7.6 [54] (options -n 1 -i 1000); convert the resulting SAM file into BAM using samtools v.1.2 [55]; trim primer sequences from reads using ivar v.1.3.1 [56] based on the coordinates inferred at step 3; extract the clipped reads and the covered part of REF_INIT (<1.5kb length) from the BAM file using bedtools v.2.29.2 [57]; Assign REF_CUR = REF_INIT; iterate steps 5-8 until no more called variants are accepted;
5. Map the clipped reads obtained at step 4 against REF_CUR using smalt;
6. Call SNPs and indel variants with lofreq v2.1.5 [58] ( -C 4 --call-indels --no-default-filter --use-orphan) and filter them, again with lofreq v2.1.5 (-Q 20 -K 20 --no-defaults -v 4 -V 0 -a 0.500001 -A 0); if no more variants are called, exit;
7. Detect and mask low coverage regions (excluding called deletions) using bedtools-2.29.2 and bedops v2.4.39 [59];
8. Apply the called variants and regions to mask to REF_CUR using bcftools-1.10.2 [55] to produce REF_NEXT; assign REF_CUR = REF_NEXT.

### Dataset preparation

To obtain data on HIV-1 diversity in Russia beyond Oryol Oblast, we downloaded the 14,365 sequences of HIV-1 collected in Russia from Genbank on 2021-08-16 (“HIV-1” AND “Russia” query). Of those, we selected the 8,560 sequences that produced an at least 800bp hit of blastn against the target *pol* fragment of at least one reference sequence from [52]. We then additionally filtered out the 1,491 sequences that were sampled outside of Russia and processed by Russian research groups thus erroneously matching the ‘Russia’ query; and the 26 samples from Oryol Oblast already in our dataset, leaving us with 7,043 samples. Genbank metadata was parsed using a custom python script for 6,252 samples; for the remaining 791 samples, fuller metadata were provided by our collaborators.

Together with the 922 sequences collected in Oryol Oblast, our Russian dataset comprised 7,965 sequences. Among the 922 Oryol samples, we identified 94 patients who had more than one sample sequenced. For each of these patients, we marked all samples except the earliest one for exclusion later in the pipeline.

### Sequence alignment and processing

Sequences were putatively aligned against the HXB2 reference *pol* region using mafft [60] (option --auto) and cropped to include only the coding part of the *pol* fragment (HXB2 coords 2252-3539). Additionally, we filtered out the sequences that either (a) had insertions relative to the HXB2 reference longer than 50bp, or (b) were shorter than 1,100 bp, leaving us with 6,356 sequences. These sequences were further aligned more accurately by the HMM-align algorithm of HIValign [61], allowing for up to 10 codons to compensate for a frameshift. From this alignment, we excluded 154 sequences carrying premature stop codons, frameshifts, or more than ten Ns (missing data characters). Sanger sequences were somewhat shorter than sequences produced by NGS; to make sequences comparably informative, we excluded codons with more than 5% of gaps or Ns from the alignment. The resulting alignment of 1,113 sites (positions 2,253 to 3,365 in the HXB2 reference) contained 6,202 sequences, including 864 samples from Oryol Oblast.

### Subtyping and DRM annotation

We used the SierraPy client [62] to assign HIV subtypes and predict drug resistance mutations. A minor fraction of samples were assigned to mixed variants (CRF02+A and A+B in Fig. 2). For further analyses, we kept only those samples corresponding to the three genetic variants most abundant in Oryol Oblast: A, B, and CRF63. Each of these three variants showed monophyly on the Russian phylogeny (Fig. 2), indicating that they can be identified robustly from a short genomic fragment. To estimate the contribution of mutations at sites associated with drug resistance (DRM), we additionally defined a set of relevant DRM sites as the 23 codons that were reported to carry DR-associated mutations in at least 5% of our samples, and repeated some of our analyses on an alignment with these sites masked (Supplementary Fig. 15,16 and Supplementary Table 5).

### Phylogenetic analyses

To validate subtyping and check the locations of samples from repeatedly sequenced patients, we reconstructed a putative Russian-wide phylogeny for the 6,202 samples using Fasttree2 [63] (double-precision release).

For the final dataset, we kept only the sequences of the earliest samples for all patients whose samples were sequenced more than once; we also excluded the three pairs of samples coming from the same patient but separated by more than 10% genetic distance (Supplementary Fig. 1), leaving us with 768 Oryol and 5,328 non-Oryol sequences. We then used Fastree2 to reconstruct phylogenies for A, B, and CRF63 variants and analysed them separately using Treetime [64] as follows. First, the trees were rerooted to maximize the temporal signal. Next, we reconstructed the ancestral nucleotide states of internal nodes in order to convert the tree to nonbinary by removing branches that did not carry any mutations. On the resulting nonbinary tree, we then determined geographical states of the internal nodes using a two-state (Oryol vs. non-Oryol) mugration model. Finally, on the basis of sequences with a known year and month of sampling, we inferred the dates of internal nodes which were used as LCA estimates (see Fig. 5), and the slope of the genetic distance vs. time regression which was used as a prior for evolutionary rate in BEAST and BEAST2 analyses (see below).

### Identification of imports

We identified imports by the depth-first search of the largest clades that (1) included at least 80% of Oryol Oblast samples, (2) had a bootstrap support of at least 0.8, and (3) had the last common ancestor that had an at least 80% posterior probability of being an Oryol node (Fig. 3). For imports resulting in just a single sequence (Oryol Oblast singletons), criteria 2 and 3 were ignored. To study the dependency of the number of inferred import lineages on the number of Oryol (Fig. 5) and non-Oryol (Fig. 6) sequences available, we randomly subsampled different numbers of Oryol or non-Oryol samples in 1,000 trials, inferring in each trial the resulting number of imports using criterion (1) above.

### Bayesian phylodynamics

In order to infer the epidemiological parameters of subtype A sub-epidemic, we used a recently developed implementation of a birth-death model in BEAST2 that jointly analyses multiple independent clades and inferres a single set of epidemiological parameters shared across all clades [12]. To our knowledge, this is the only implementation that is capable of aggregating clades spanning different time intervals and setting any arbitrary timepoints where epidemiological parameters can change in a piecewise-constant fashion, allowing us to use priors informed by data. As birth-death models require at least one parameter to be fixed or defined in a narrow range, we put a strict prior on sampling proportion. To obtain it, we selected samples that were sequenced soon (in less than a year) after the initial diagnosis which we denote as Dataset I. In Dataset I, most samples were sequenced in 2018 and 2019, eight samples were sequenced in 2014-2017, and none before 2014. We thus put a four dimensional prior on the sampling proportion which was allowed to change on the first day of 2014, 2018 and 2019, with the mean equal to the number of samples sequenced in a time interval divided by the number of new diagnoses in this interval based on the reported case counts (Supplementary Table 1). We then used these priors on both Dataset I and Dataset II composed of all samples. In contrast to the sampling proportion, the reproduction number and the rate of becoming uninfectious were assumed to be time-independent and thus unidimensional. We used a relaxed lognormal uncorrelated clock with ulcd.mean and ulcd.std shared across all imports. For ulcd.mean prior, we used the normal distribution with mean equal to the regression slope inferred by Treetime and sigma equal to 0.001. All other priors were kept default. Priors used in the multi-tree birth-death analysis are summarized in Supplementary Table 2. The analysis was run for 250 million steps; we discarded the first 10% steps as burn-in.

We used the same set of priors to infer the epidemiological parameters of the largest clades of variants A and CRF63. The parameters were inferred using the BDSKY [65] package of BEAST2 [66]. The skylinetools package [67] was used to define appropriate time intervals for sampling proportion. Priors used in this analysis are summarized in Supplementary Table 3. The analysis was run for 100 million steps; we discarded the first 10% steps as burn-in.

The logistic growth dynamics for the same two clades was inferred in BEAST v1.10.4 [68]. Priors are provided in Supplementary Table4. The analysis was run for 100 million steps; we discarded the first 10% steps as burn-in.

Convergence of the produced MCMC trajectories was assessed using Tracer [69].

### EpiEstim

We used the EpiEstim package [70] implemented in R to infer the dynamics of R_e_ from the reported incidence data. We constructed the distribution of serial intervals based on the estimates of HIV-1 transmission rates at different stages of infection inferred by Hollingsworth et al. in [45]. As our incidence data is provided per year, we converted the reported transmission rates to be year-wise (see Supplementary Fig. 14). The dynamics of R_e_ was inferred using a five-year sliding window.

### Analysis of transmission lineages

We tested whether the two categories of samples, male gender and IDU route of transmission, are overrepresented in transmission lineages compared to singletons. For this purpose, we reshuffled the gender or transmission route labels of Oryol Oblast samples 10,000 times and obtained the distribution of the expected number of samples of the tested category belonging to transmission lineages, and the expected number of transmission lineages carrying such samples. Additionally, we used https://github.com/appliedmicrobiologyresearch/Influenza-2016-2017 to test co-occurence of samples from each of the two tested categories within the same lineage.

To test whether transmission lineages are preferably seeded by IDUs, we constructed two match-paired datasets. First, we sorted all transmission lineages by the earliest diagnosis in the lineage. Then, in this sorted list, we marked lineages as IDU-founded and HET-founded based on the earliest sample and selected time-matched pairs of IDU- and HET-founded lineages such that in a consecutive row of lineages with the same founder type, the earliest lineage was taken; this ensured that IDU-founded lineages could not be overrepresented due to the population structure being mostly comprised of IDUs in 1990s. We compared the time between the date of the LCA and the date of the first diagnosis in a lineage using the paired Wilcoxon test.

Second, we sorted all lineages by the median date of diagnosis, selected the ones with both HET and IDU samples, and compared the dates of diagnosis of the earliest HET in odd lineages and the earliest IDU in even lineages, again using the paired Wilcoxon test.

## Data availability

Partial sequences of the HIV *pol* gene produced in this study are being deposited to GenBank; metadata will be made available as a Supplementary Data file after Genbank IDs are assigned.

## Data Availability

Partial sequences of the HIV pol gene produced in this study are being deposited to GenBank; metadata will be made available as a Supplementary Data file after Genbank IDs are assigned.

## Acknowledgements

We thank all patients who contributed to this study. This work was funded by the Russian Science Support Foundation (project no. 21-74-20160 to G.A.B.).

## Author contributions

YS collected epidemiological, clinical and demographic data. NE enrolled patients into the study. EB collected clinical samples. AK performed HIV-1 genome fragment amplification. DS and AK performed sequencing. KRS performed consensus calling and bioinformatic analysis. DK conceived the project. DK and GAB designed the project. DK, VP, and GAB supervised the project. KRS, DK, and GAB wrote the manuscript.

## Supplementary Figures

**Supplementary Figure 1.**
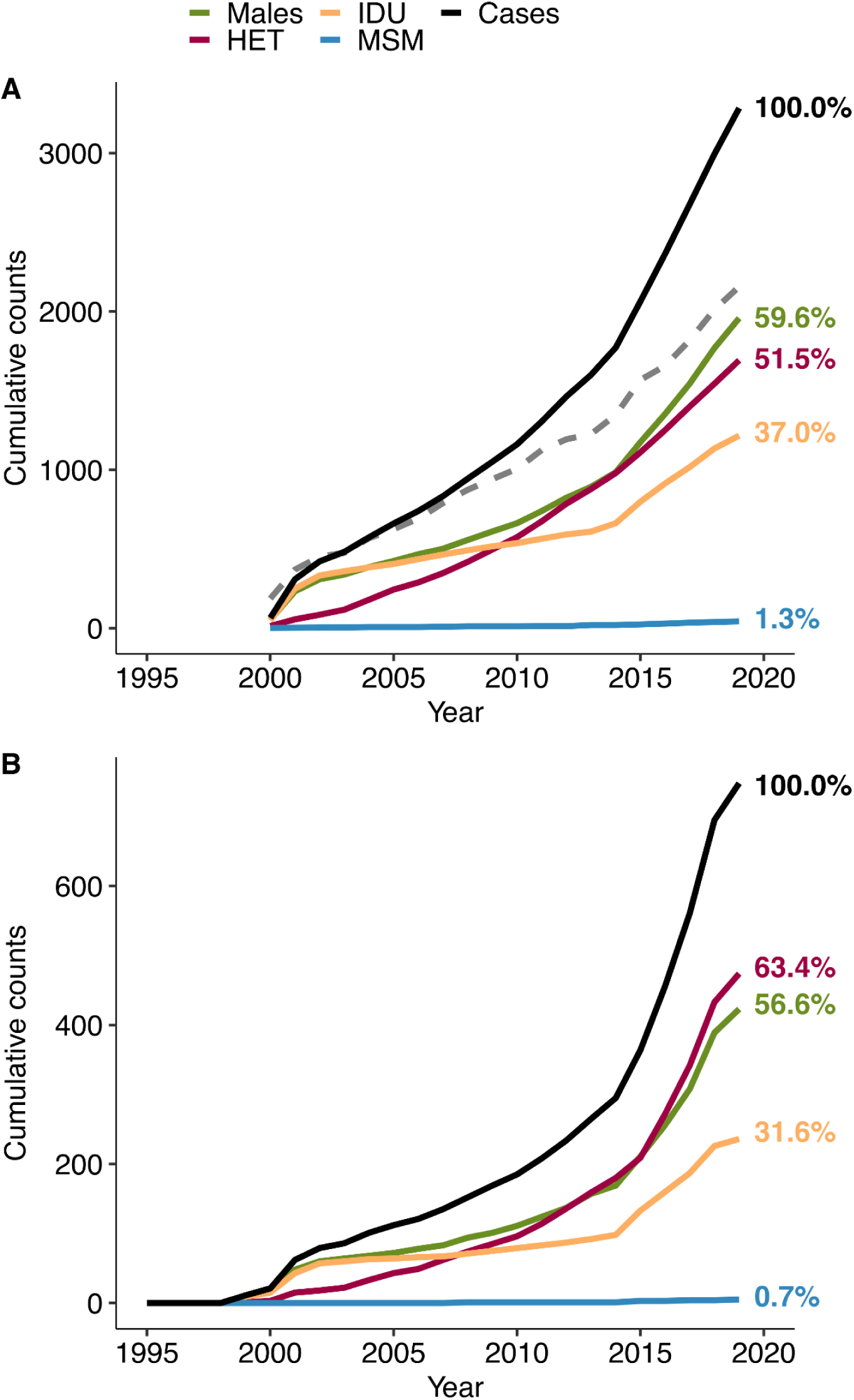
The statistics inferred from the sequenced dataset agrees with the official statistics on the Oryol Oblast. The plots show the cumulative number of different categories according to (A) the statistics provided by the Oryol Oblast AIDS center and (B) sequenced samples analysed in this study. Dashed gray line on A shows the total number of HIV-1 positive people registered in Oryol Oblast every year and differs from black line due to deaths and migrations.

**Supplementary Figure 2.**
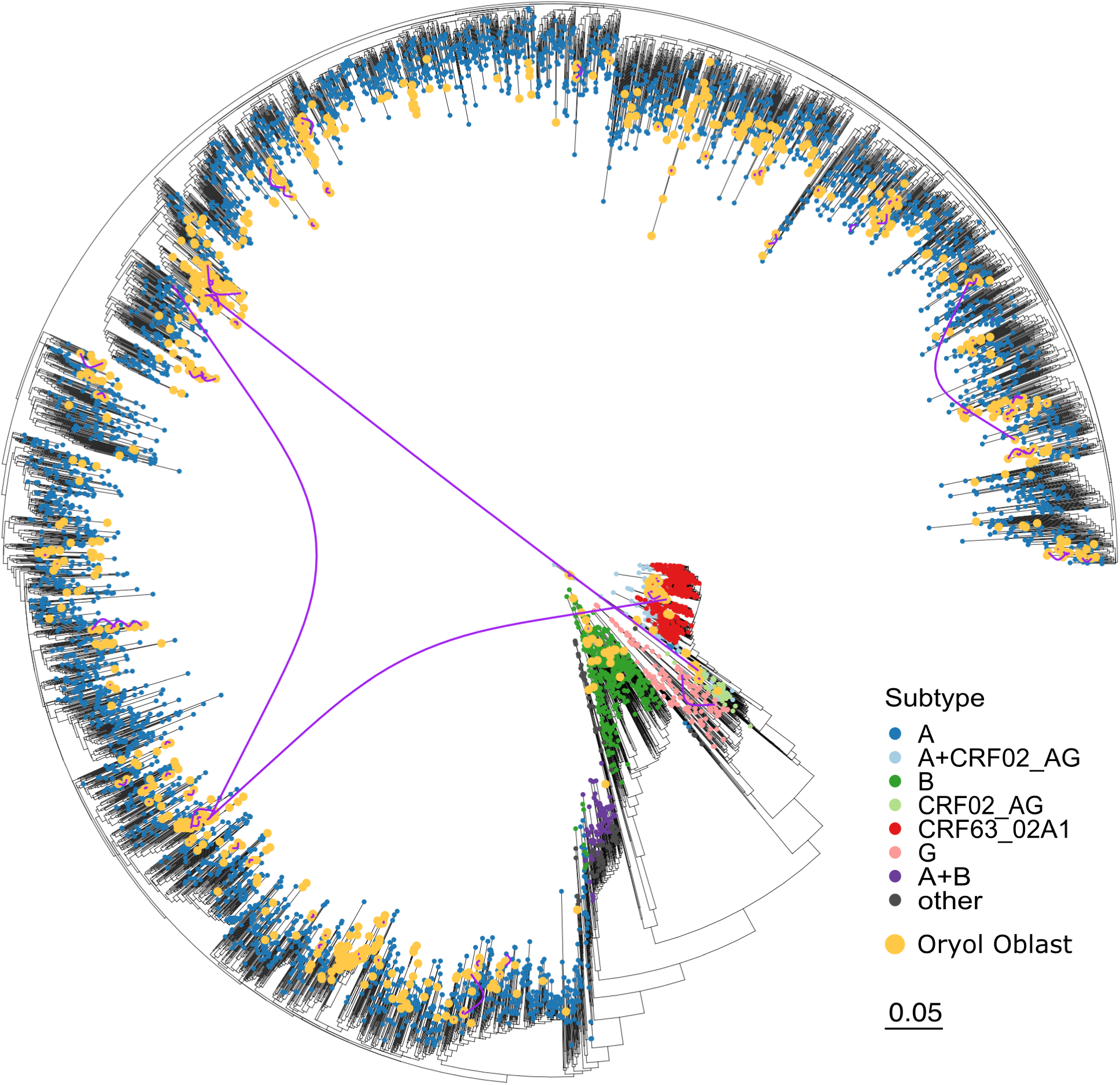
The complete phylogenetic tree of the combined Russian dataset, including multiple samples from repeatedly sequenced patients (joined with purple arcs).

**Supplementary Figure 3.**
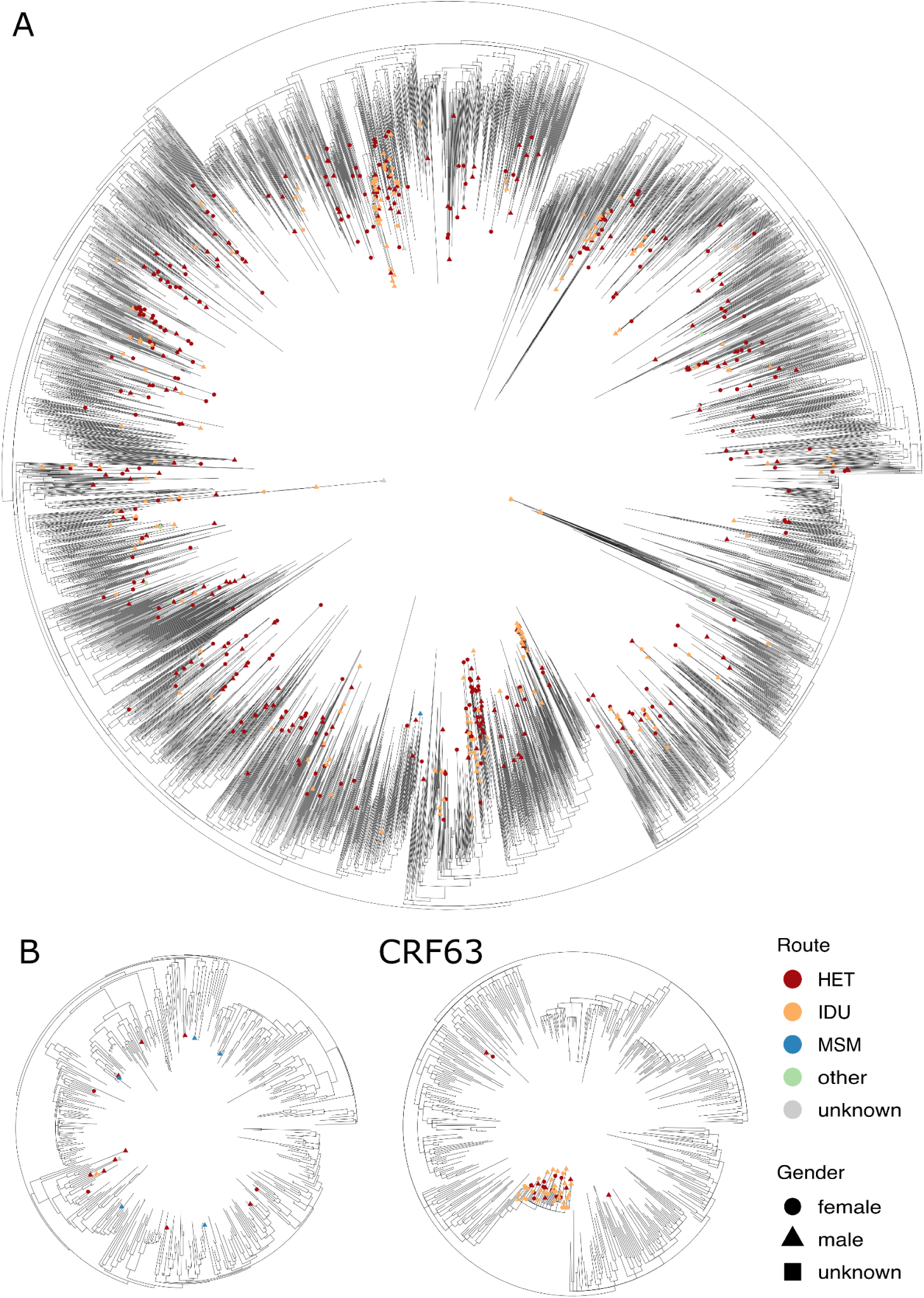
Phylogenetic trees reconstructed for subtypes A and B and CRF63. When more than one sample per patient was available, only the earliest sample was used.

**Supplementary Figure 4.**
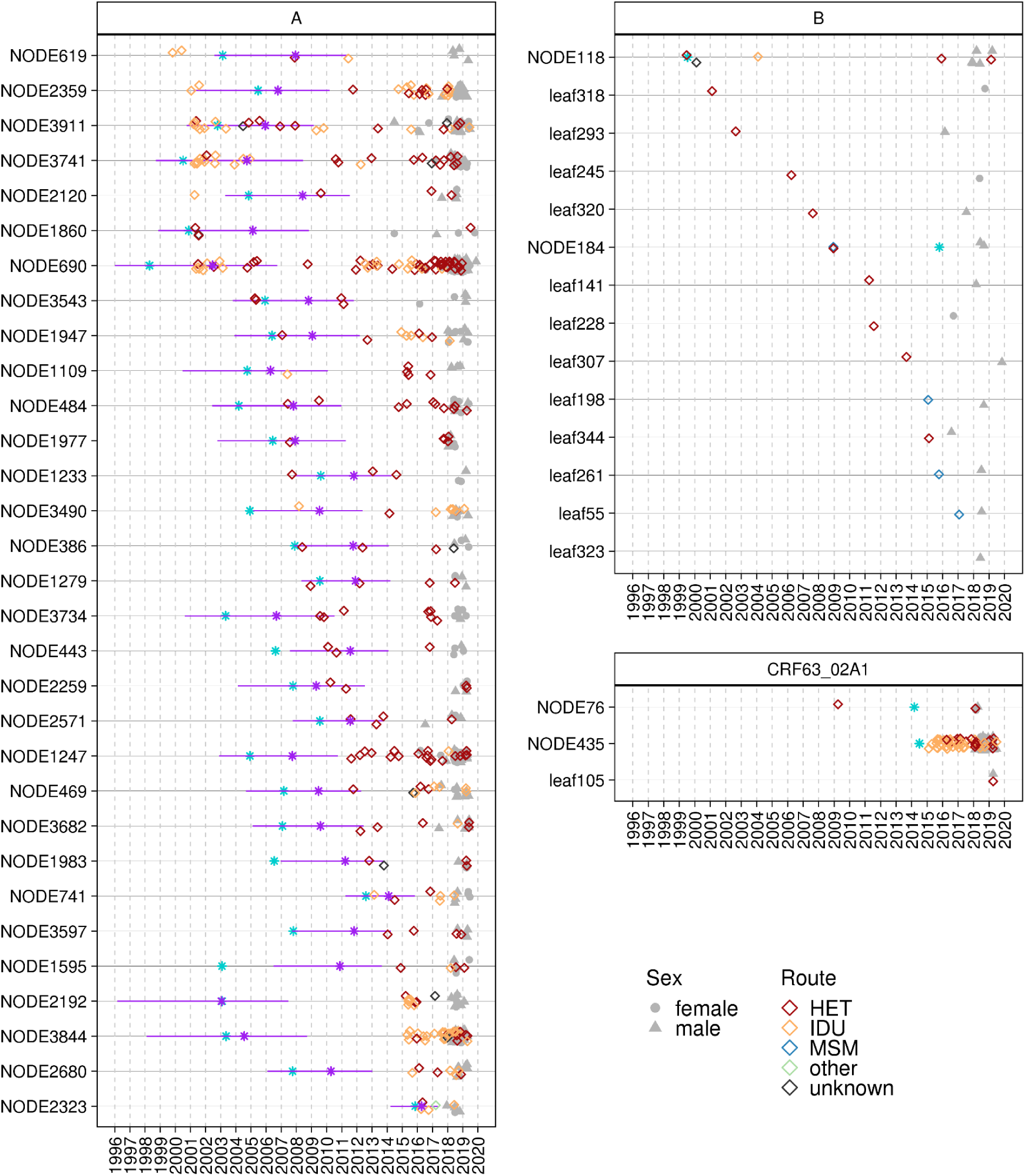
Introductions sorted by the earliest diagnosis.

**Supplementary Figure 5.**
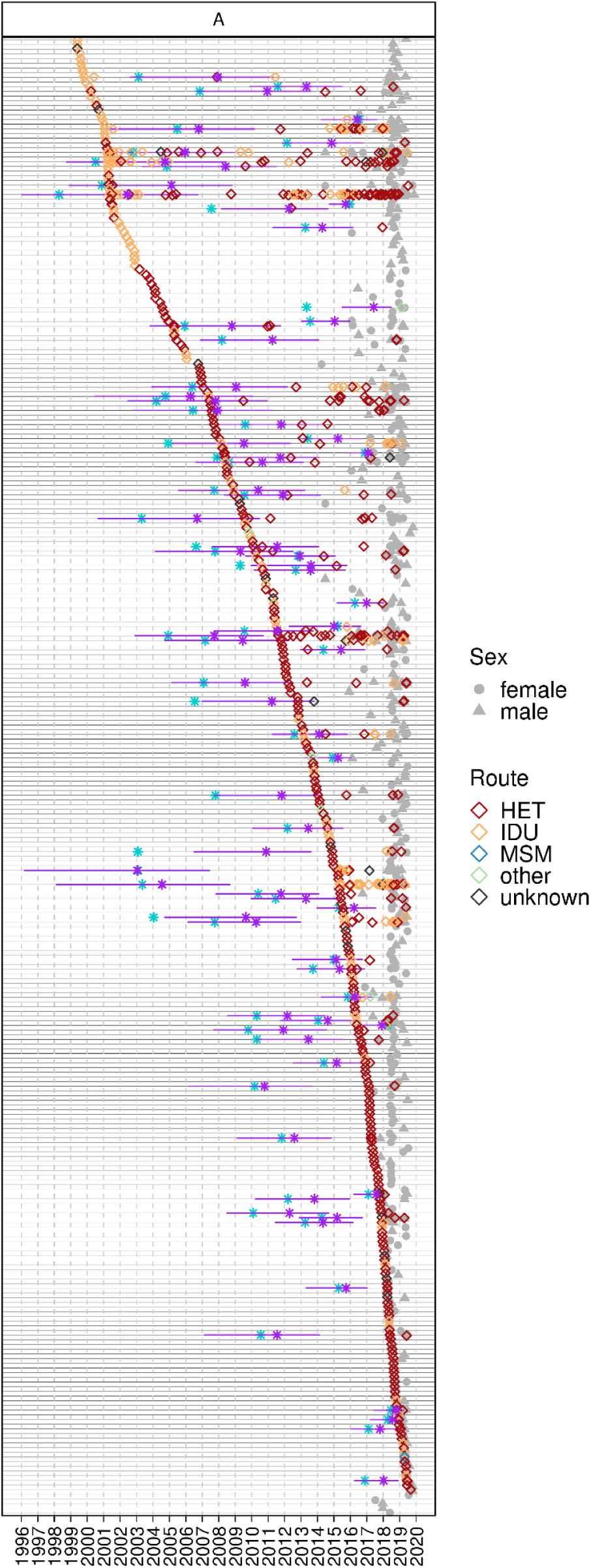
All subtype A introductions sorted by the earliest diagnosis. Legend as in Fig. 5.

**Supplementary Figure 6.**
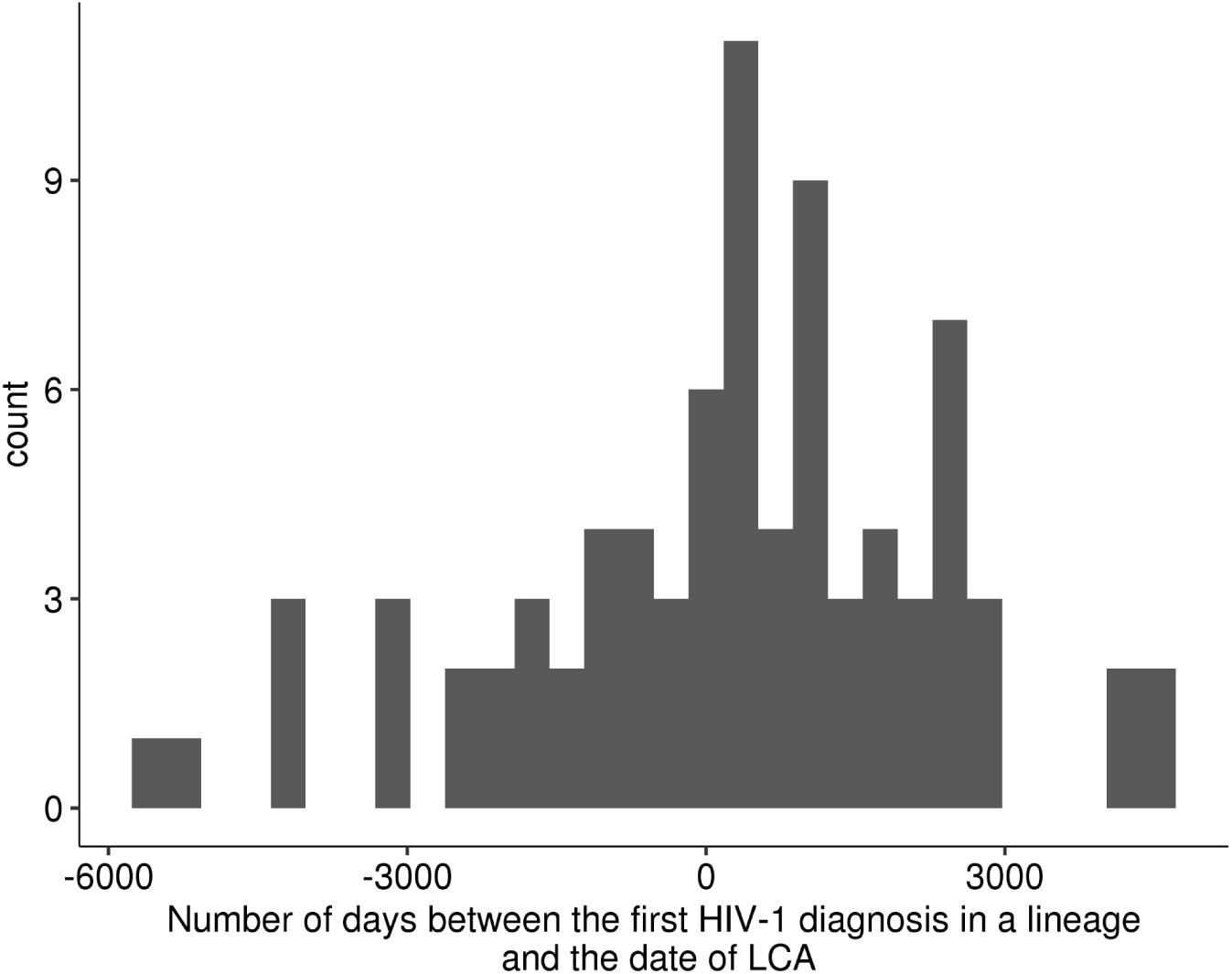
The number of days between the earliest HIV-1 diagnosis in a lineage and the inferred date of its LCA.

**Supplementary Figure 7.**
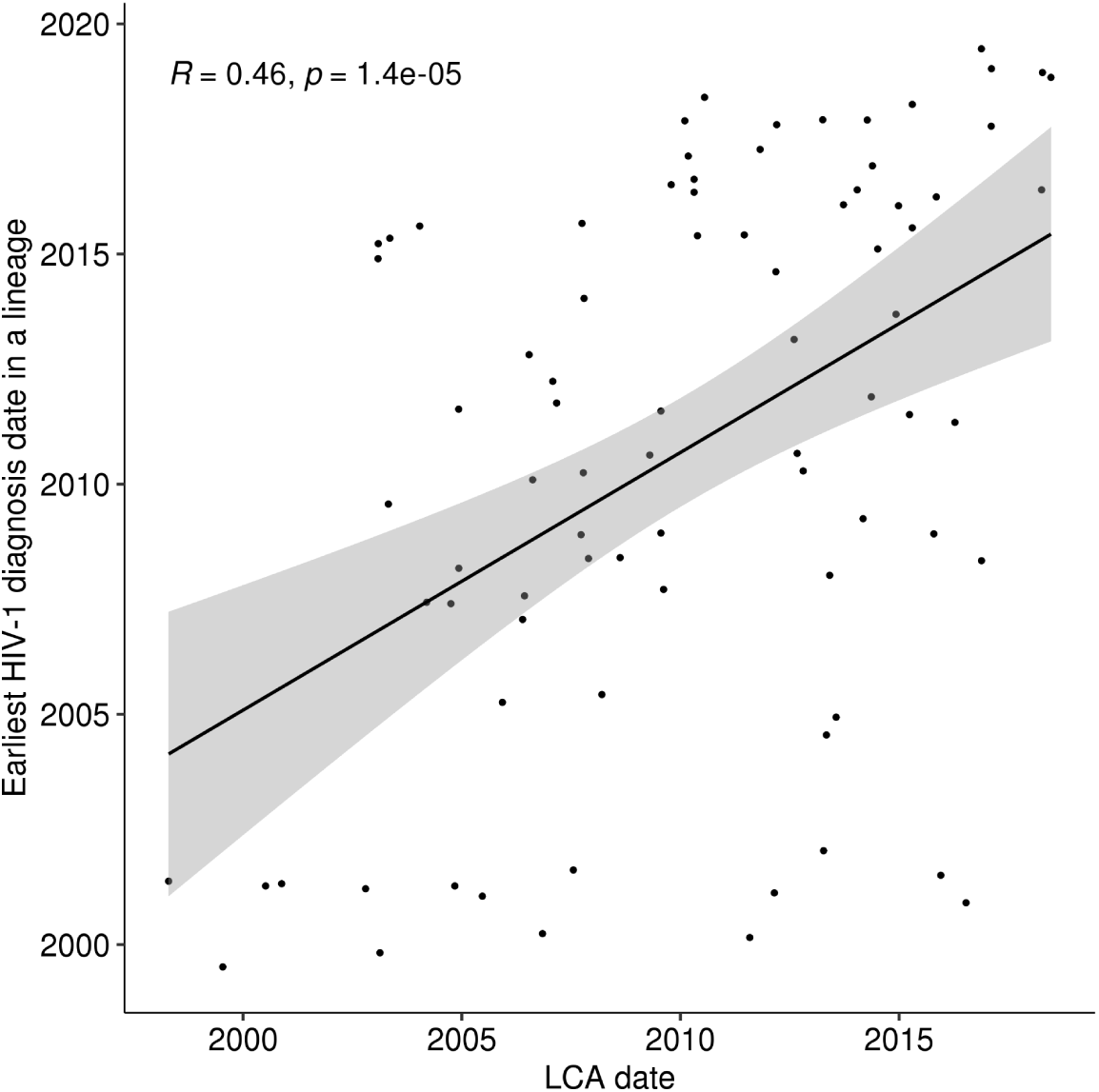
The inferred LCA date correlates with the date of the earliest diagnosis in a lineage.

**Supplementary Figure 8.**
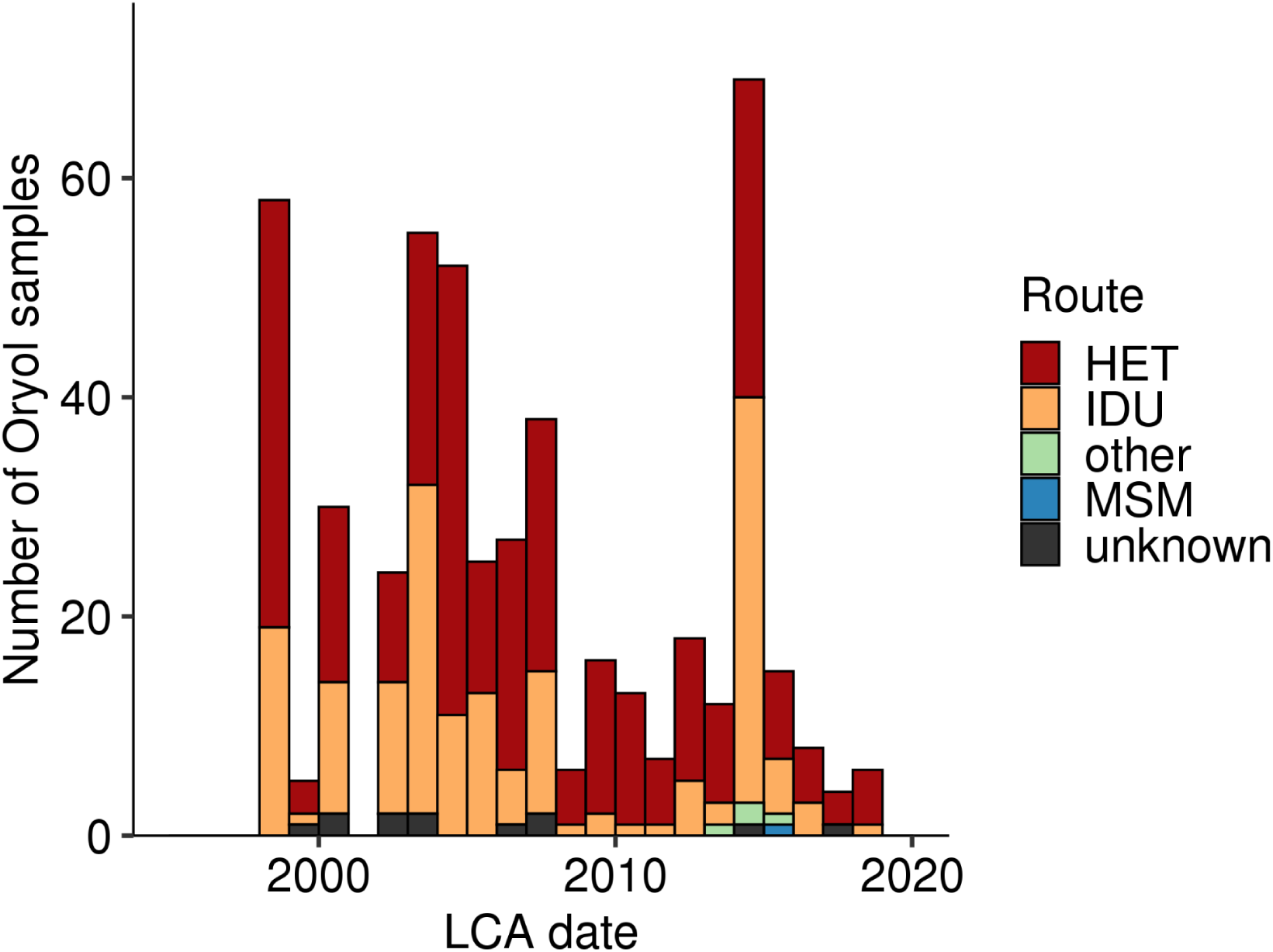
The distribution of samples by routes and the LCA date of their lineages.

**Supplementary Figure 9.**
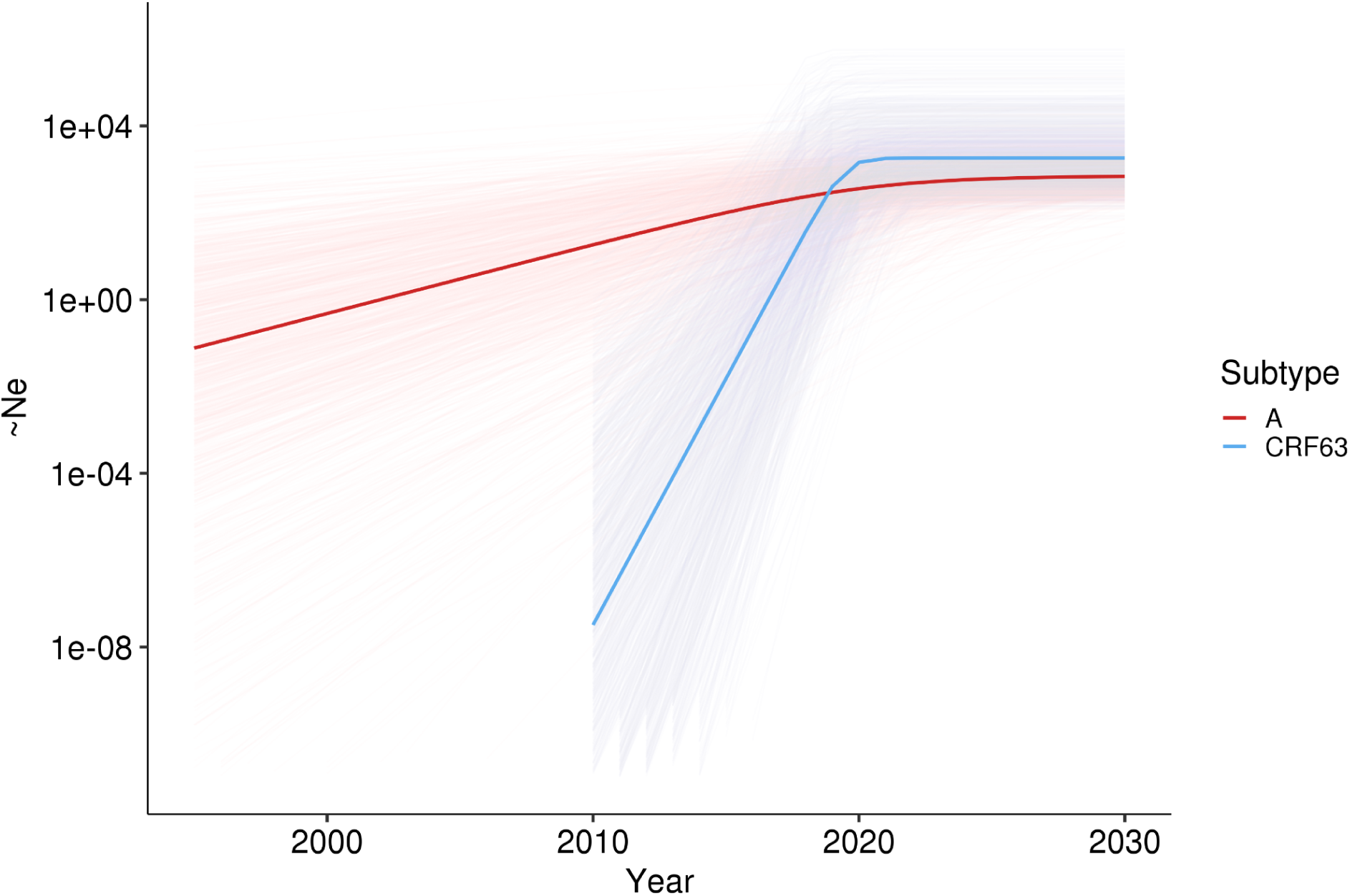
The logistic growth dynamics inferred in BEAST for the two largest transmission lineages. Shading corresponds to a 1,000 randomly sampled trajectories during the MCMC run.

**Supplementary Figure 10.**
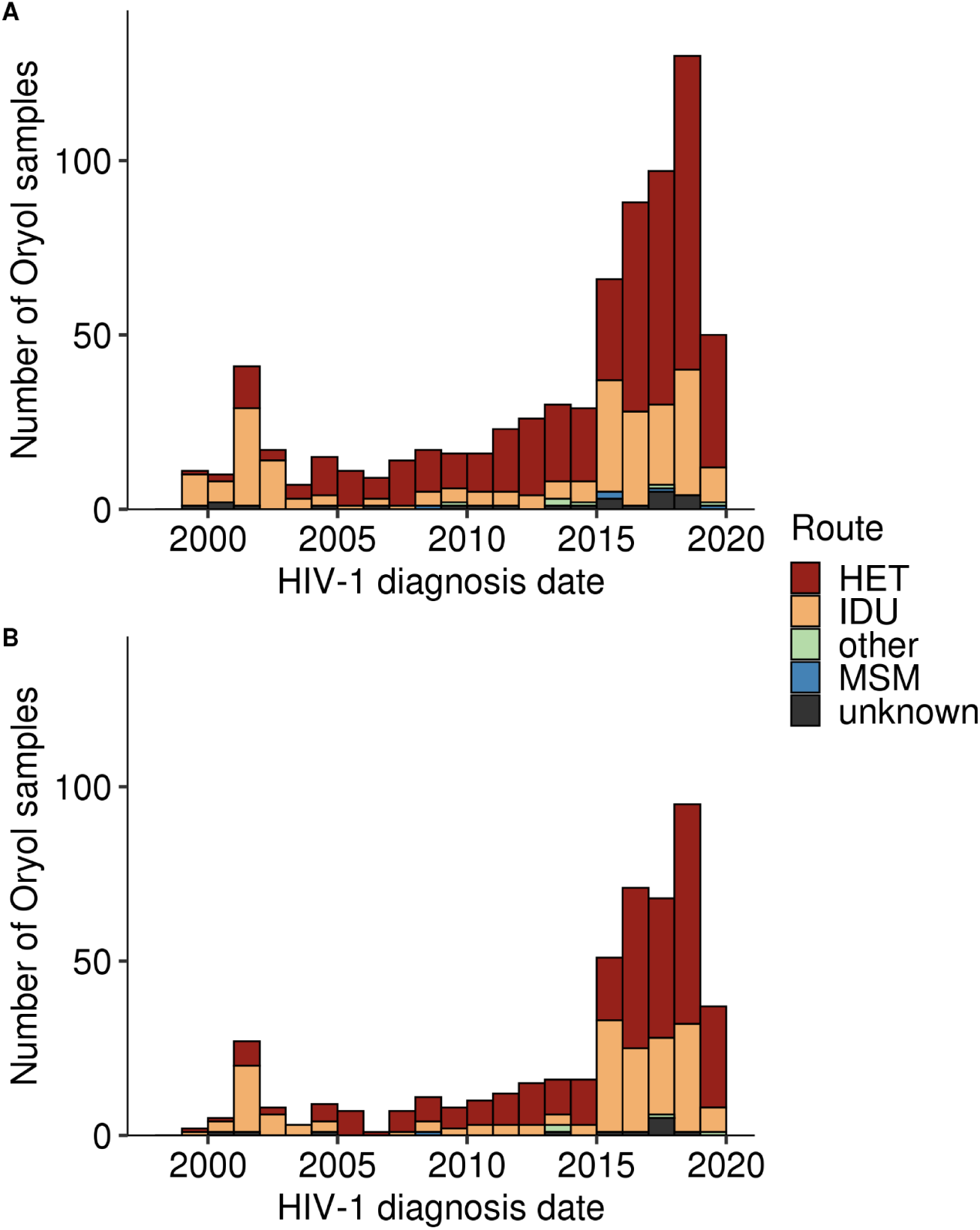
The distribution of samples by route and diagnosis date for all samples (A) and for samples belonging to transmission lineages (B).

**Supplementary Figure 11.**
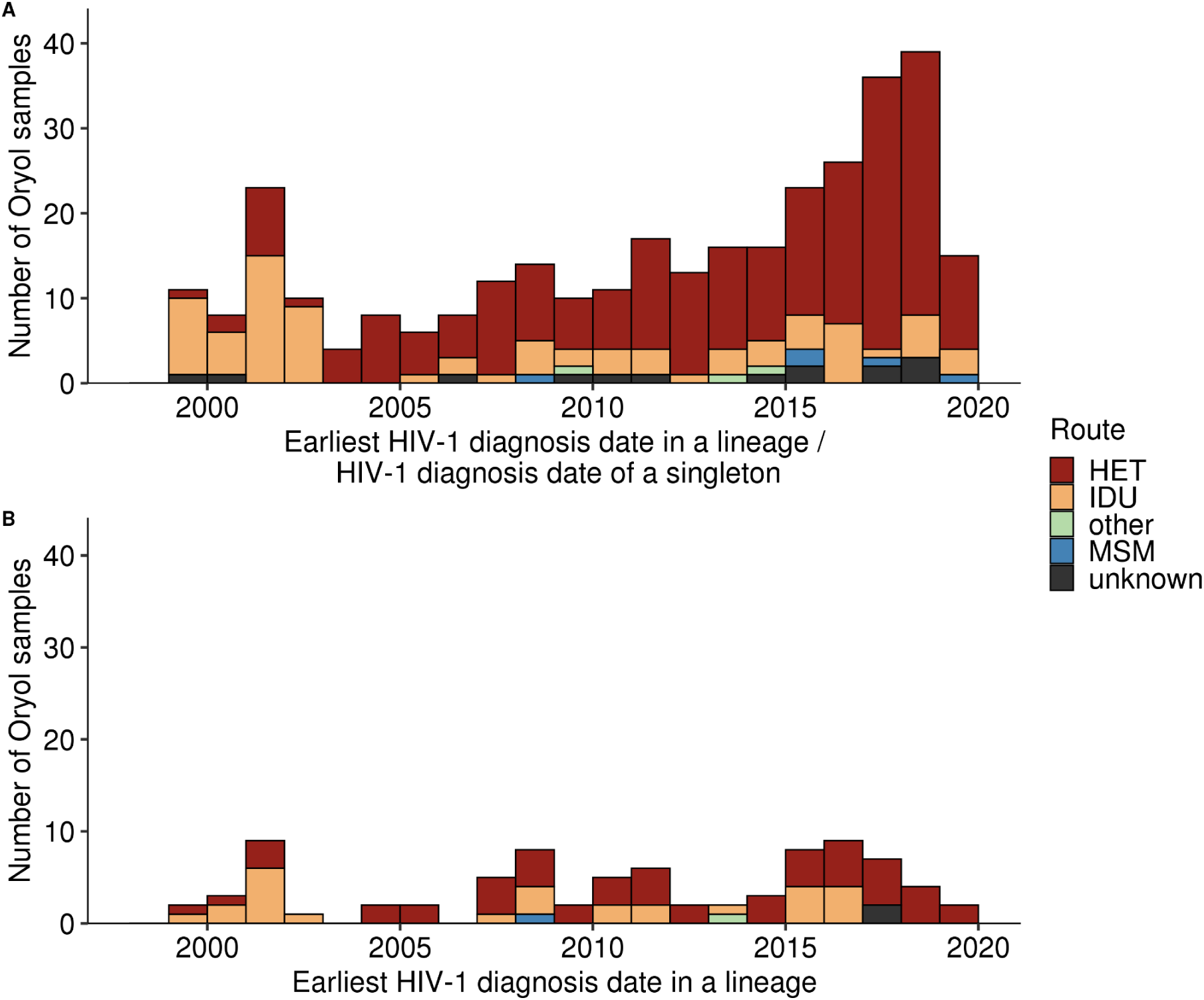
The distribution of the earliest sample per import by route and the first diagnosis date for all imports (A) and transmission lineages (B).

**Supplementary Figure 12.**
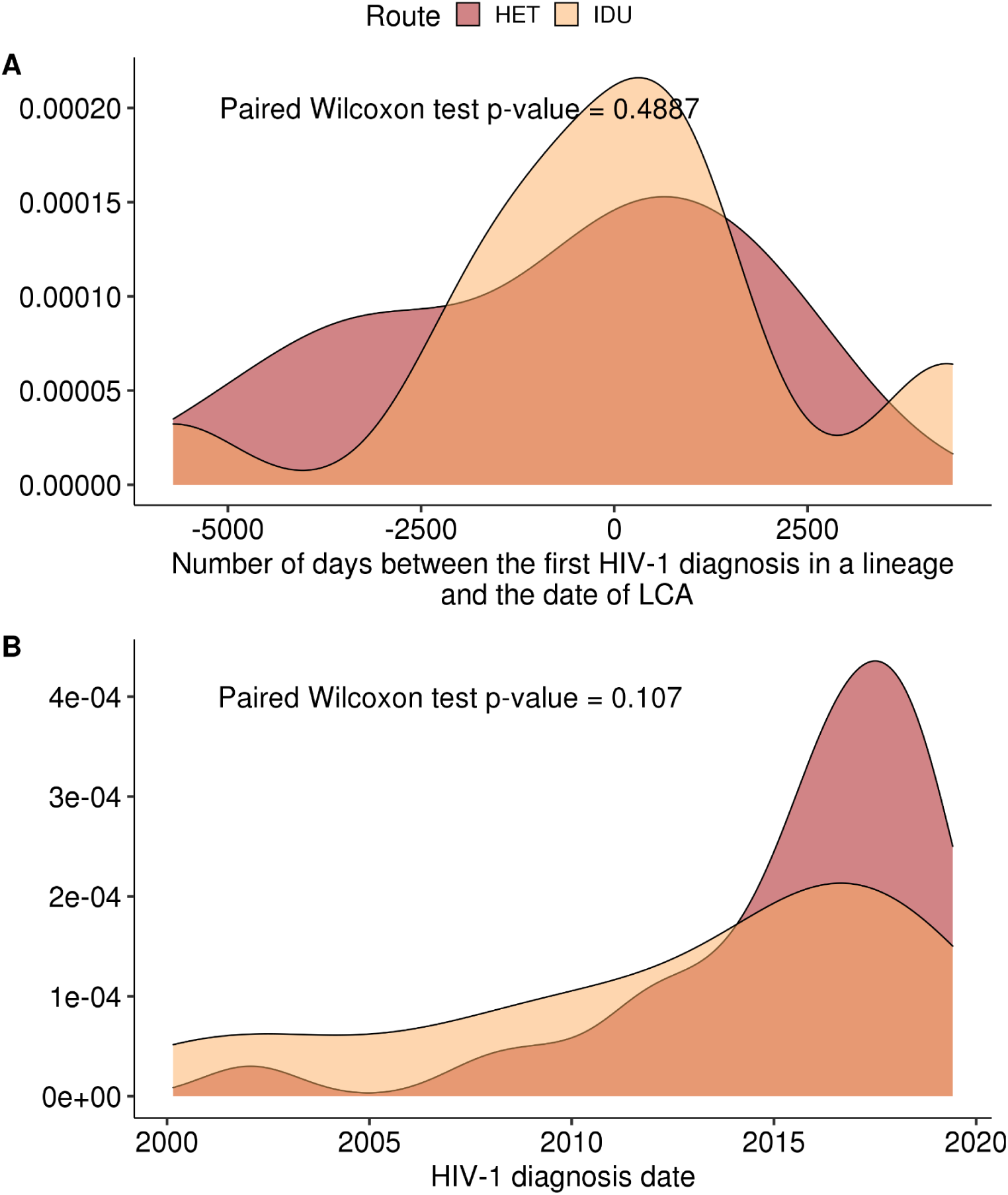
IDUs are not more likely to found a transmission lineage. A. The number of days between the first HIV-1 diagnosis in a lineage and the date of LCA in a matched-pair dataset sorted by the earliest diagnosis in a lineage (see Methods). B. HIV-1 diagnosis date of HETs and IDUs in a matched-pair dataset sorted by the median date of diagnosis in a lineage.

**Supplementary Figure 13.**
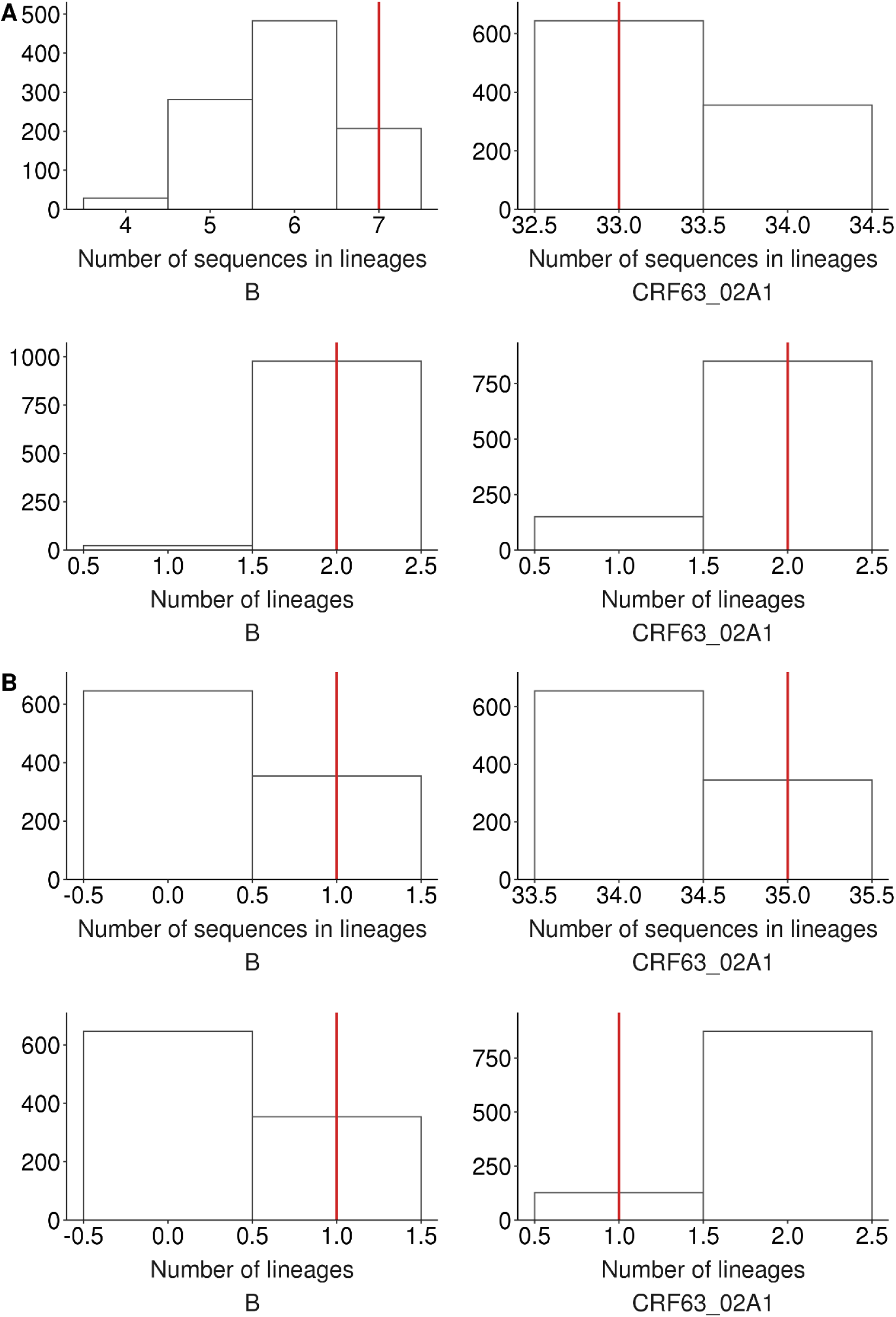
The expected distribution of the number of clustered sequences and the number of clusters carrying (A) males or (B) IDUs within subtypes B (left) and CRF63 (right).

**Supplementary Figure 14.**
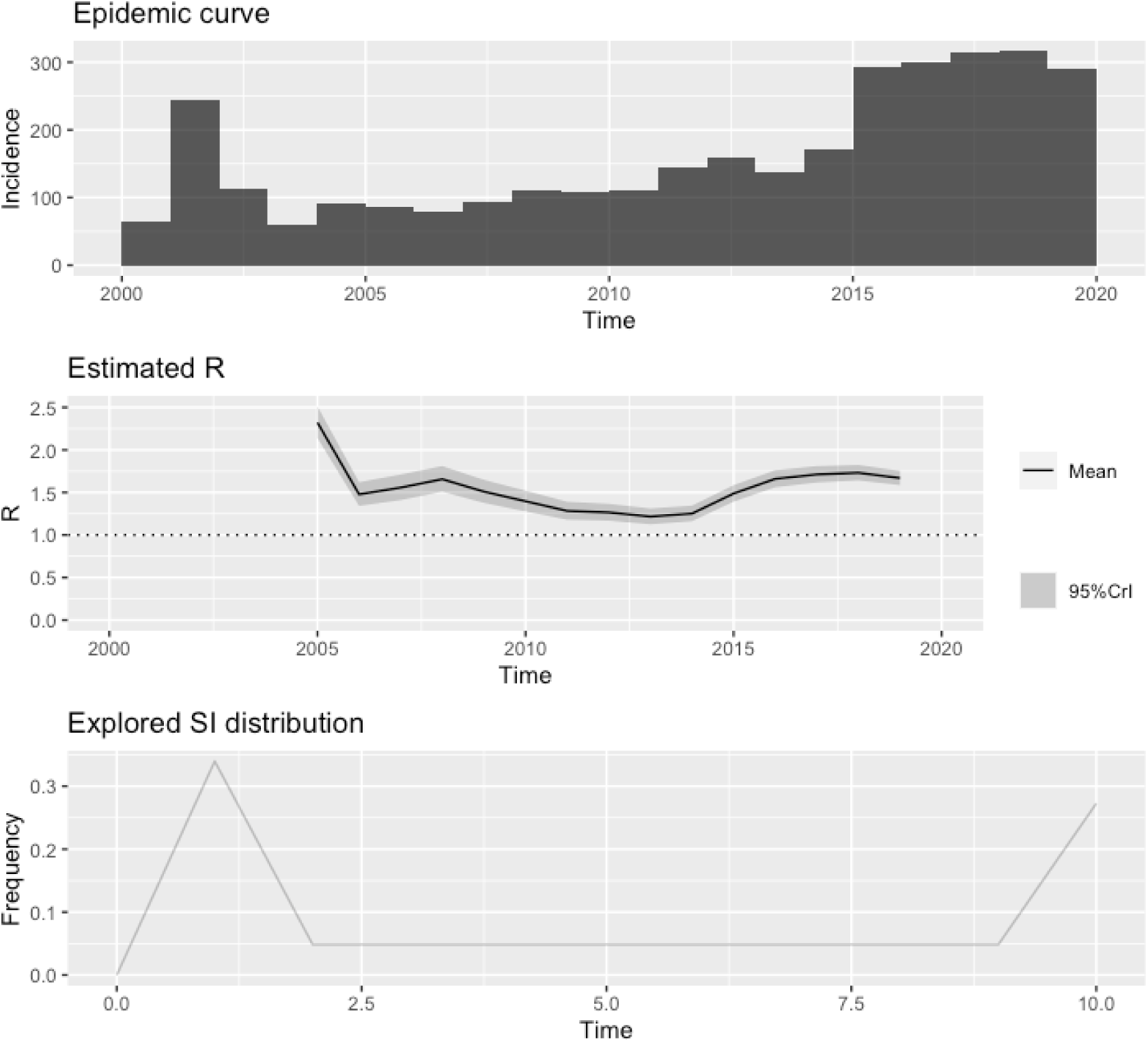
The reproduction number dynamics inferred by EpiEstim on the basis of case counts.

**Supplementary Figure 15.**
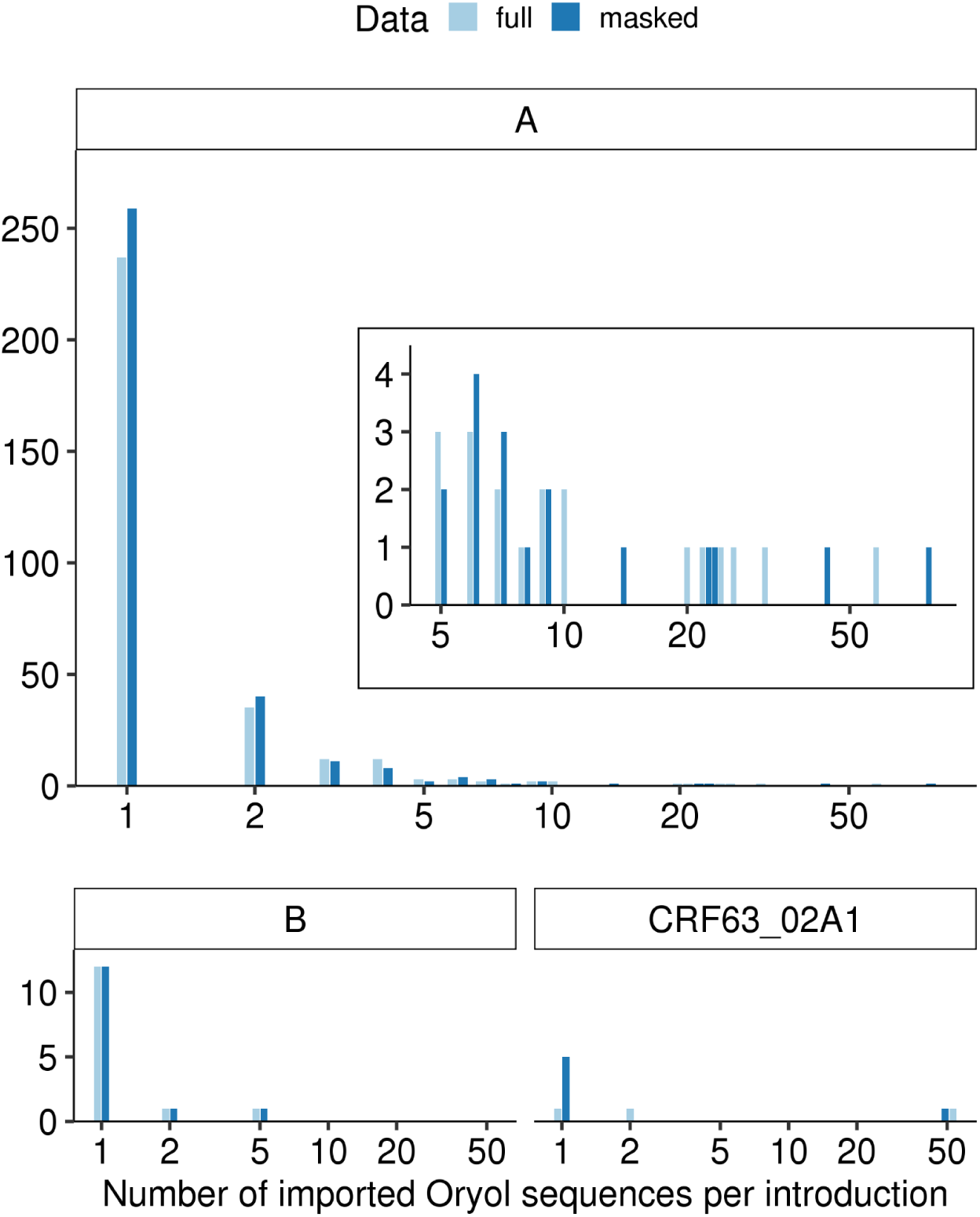
Dependence of distribution of samples across transmission lineages on DRM masking.

**Supplementary Figure 16.**
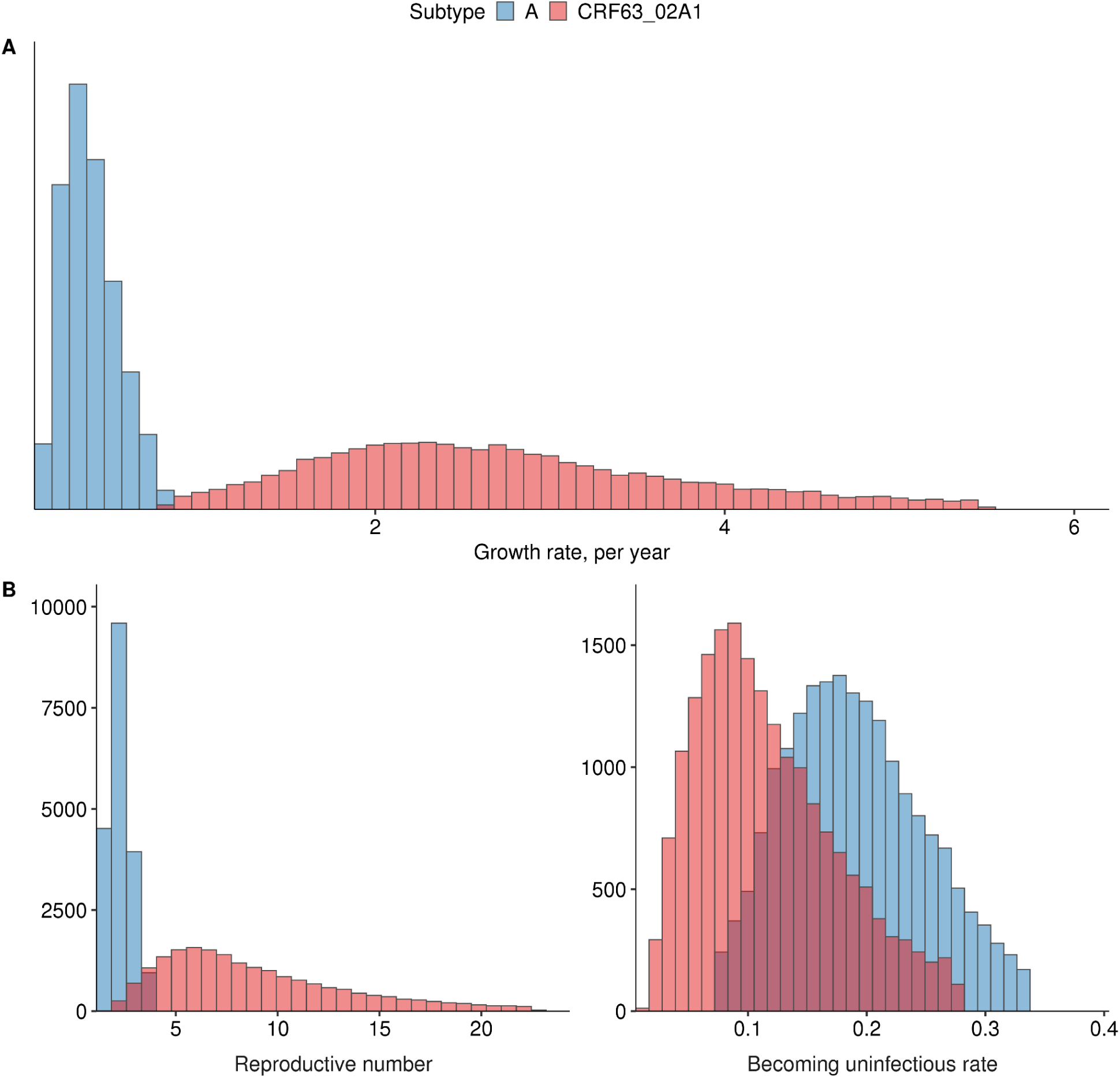
Comparison of two clades with DRM masked. A. Logistic growth, B. BDSKY

## Supplementary Tables

**Supplementary Table 1.** Inferring prior on sampling proportion. Estimated fractions are used as mean values of prior distributions on sampling proportion.

| Time period | Number of new cases | Number of recently diagnosed sequences | Fraction |
| --- | --- | --- | --- |
| Before 2014 | 1600 | 0 | 0.0001<br>(pseudocounted) |
| 2014-2017 | 1078 | 8 | 0.007 |
| 2018 | 316 | 149 | 0.473 |
| 2019 | 290 | 71 | 0.246 |

**Supplementary Table 2.**
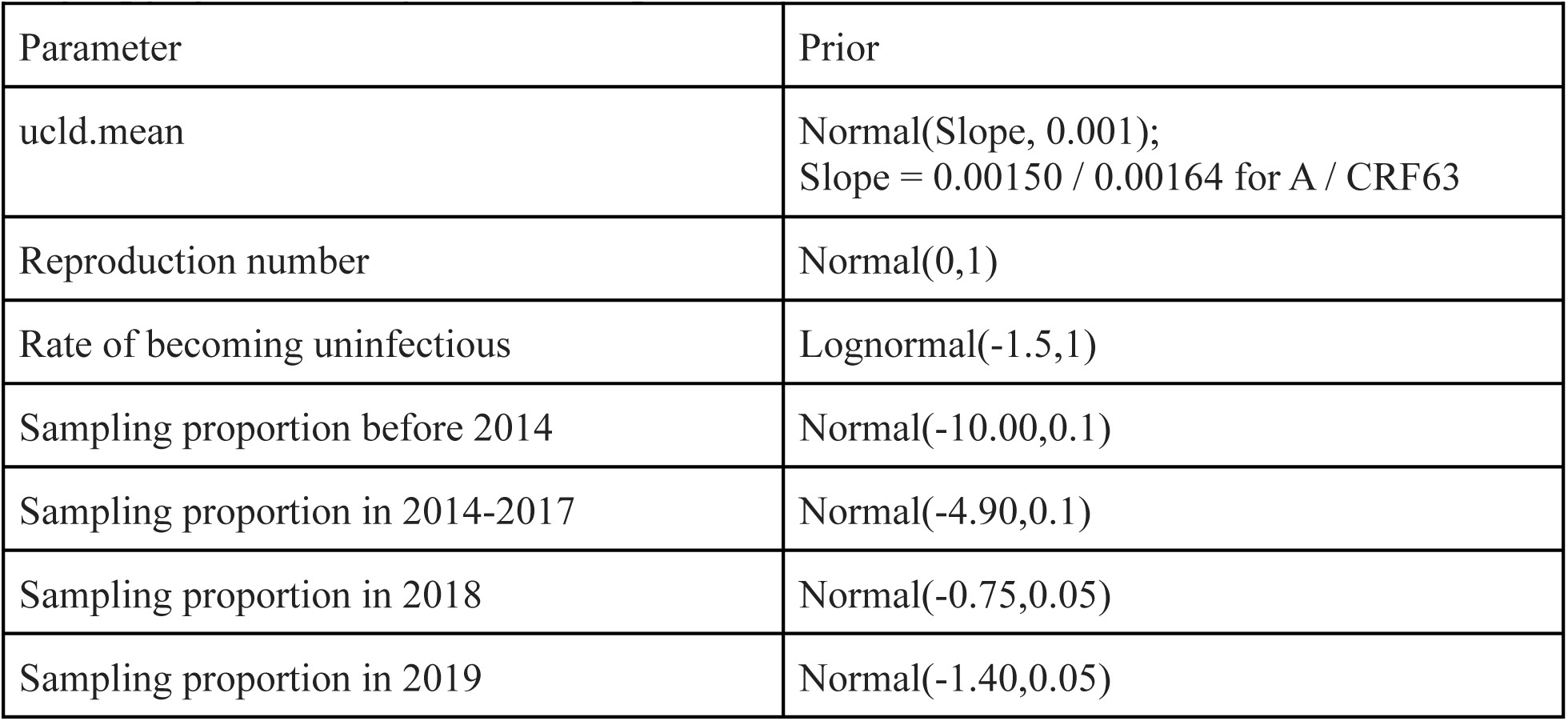
Priors used for multi-tree birth-death analysis. Reproduction number and sampling proportion are implemented in log-scale in this custom BEAST2 release.

**Supplementary Table 3.**
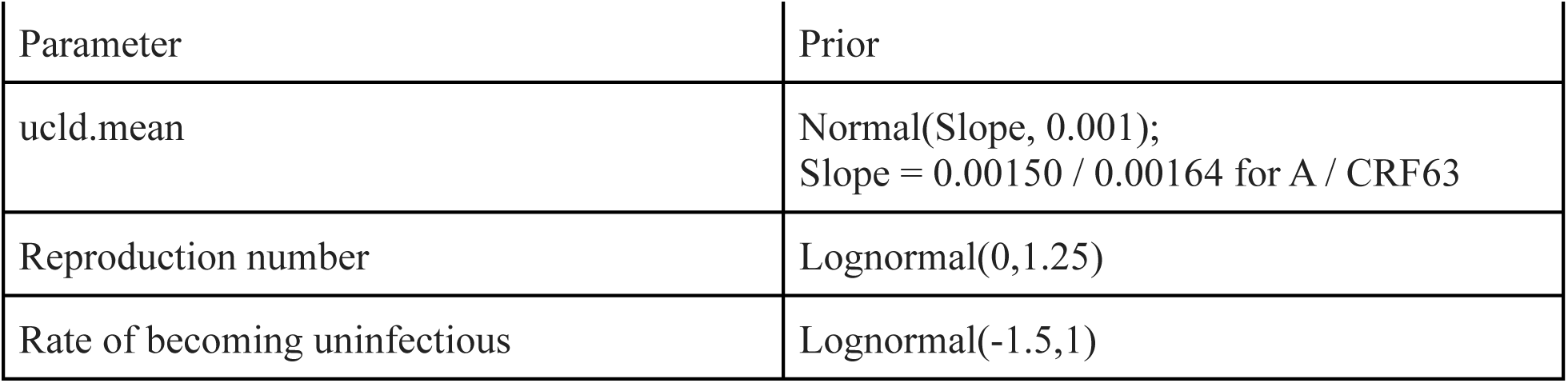
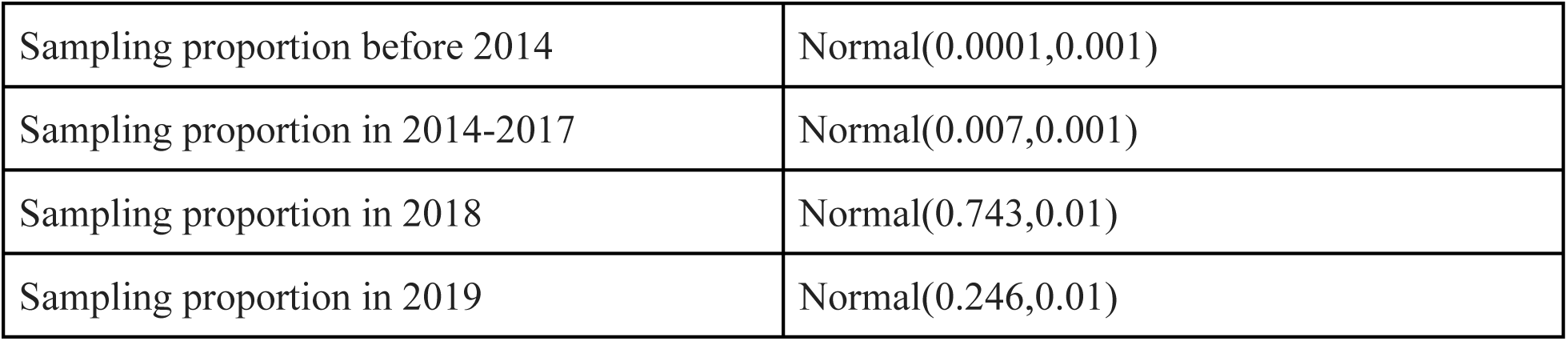
Priors used for BDSKY analysis of two clades

**Supplementary Table 4.**
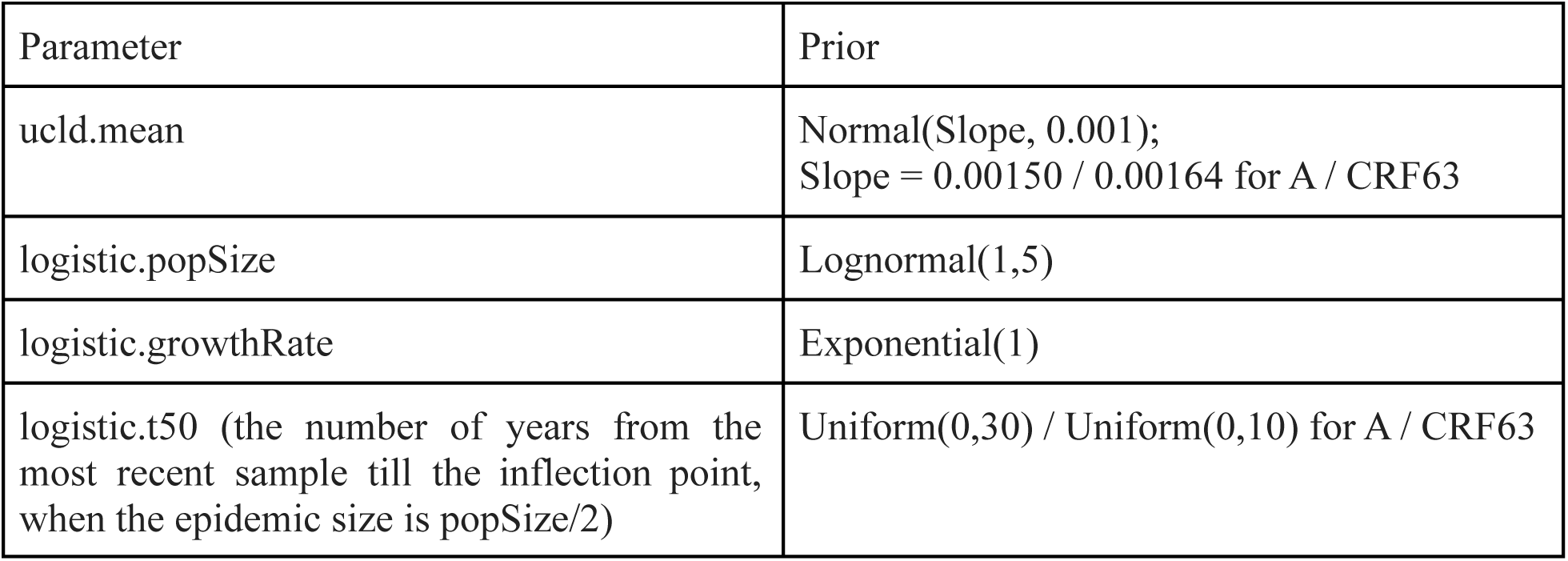
Priors used for logistic growth analysis

**Supplementary Table 5.**
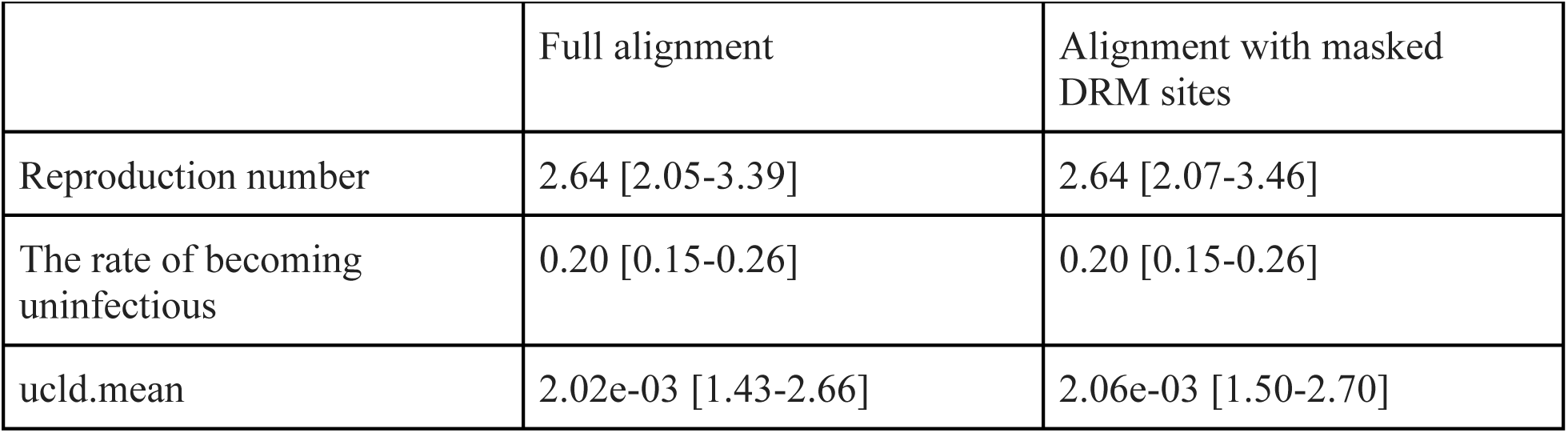
Multitree birth-death estimates produced for subtype A. Median (95% HPD) values are provided.

